# Characterizing Trainee Workload Using Paired Daily Surveys and EHR Use Data: A Mixed-Methods Pilot Study

**DOI:** 10.64898/2026.04.16.26351062

**Authors:** Karan Rai, Nicholas Bianchina, Cristina Fischer, Jessica Clawson, Lauren McBeth, Emily Gottenborg, Angela Keniston, Marisha Burden

## Abstract

**Purpose:** High clinical workload is associated with worse patient and hospital outcomes and is a well-established driver of clinician burnout. Trainees may be particularly exposed, shouldering both clinical and educational responsibilities. Evidence-based work design offers a data-driven approach to healthcare work but relies on robust workload measurements. Trainee workload remains poorly characterized, as commonly used metrics (e.g., duty hours, patient census) overlook cognitive and contextual dimensions. This pilot evaluated the feasibility of combining survey-based and electronic health record (EHR) data to characterize internal medicine (IM) trainees’ workload.

**Methods:** A pilot study was conducted including IM and Medicine-Pediatrics residents (postgraduate years 1-4) between March 31 and June 22, 2025. Participants completed daily surveys during a seven-day inpatient schedule assessing workload and work experience domains, including environment, professional fulfillment, psychological safety, autonomy, and rounding experience, using validated instruments where available. Concurrently, EHR data captured chart review, documentation, orders, and secure messaging activity. Associations between survey and EHR data were assessed.

**Results:** Among 37 eligible residents, 28 (76%) participated in the pilot capturing 166 shifts. Trainees spent 4.4 ± 1.6 (mean ± SD) minutes completing daily surveys and 8.6 ± 2.3 minutes completing the final survey. Trainees reported working 11.6 ± 1.0 hours/day and a median census of 9.0 (IQR 6.0-11.0). NASA-TLX score was 50.8 ± 12.6. Positive shift ratings were associated with lower NASA-TLX scores and perceived rounding length. First-to-last EHR login duration was 15 ± 2 hours/day, and EHR data showed 204 ± 46 active minutes/day. Login duration correlated with self-reported hours (r=0.43, p<0.0001), and notes signed correlated with self-reported team (r=0.19, p=0.013) and personal census (r=0.34, p<0.0001).

**Conclusions:** Integrating survey-based and EHR-derived workload measures provides multidimensional insight into trainee work. This novel approach supports scalable measurement and evidence-based work design interventions to improve trainee well-being, education, and clinical efficiency.

## Introduction

High clinical workload is associated with worse clinical and hospital outcomes^1,2^ and contributes to clinician burnout.^3^ Trainees, who work up to an average of 80 hours per week, are at particular risk, with nearly half of physicians-in-training reporting burnout,^4^ underscoring longstanding concern about the intensity and structure of trainee work. Clinician burnout is linked to personal morbidity, such as poor health and increased suicide risk,^5^ as well as downstream effects on patient care such as higher rates of medical errors, lower patient satisfaction, and billions of dollars in societal cost through lost productivity.^3,6^ In recent years, trainees have increasingly turned to organizing efforts (i.e., unionization)^7^ to advocate for improved work conditions and workplace protections.^8,9^ While these efforts have demonstrated promising early results in securing benefits such as vacation and stipends, collective action alone does not ensure improvement in the design of day-to-day clinical work, educational quality, or lived work experience.^10,11^

These limitations highlight the need for complementary, system-level strategies that directly address how clinical work is structured. Evidence-based work design has emerged as a promising framework that systematically applies data and human factors principles to optimize staffing, task allocation, and workflows to support patient safety, operational performance, and clinician wellbeing.^12–14^ Rather than relying solely on advocacy or regulation, this framework seeks to redesign work itself and offers a potential path to improve both operational efficiency and trainee experience.^15^ Central to this approach is the availability of valid, actionable measures of workload and work environments. Traditional metrics, such as duty hours and patient census, remain widely used in graduate medical education, but they are increasingly believed to be insufficient alone, missing critical elements of patient acuity, cognitive and emotional burden, and context that shape how workload is experienced.^12,13,16,17^ While duty hour restrictions have historically been the major focus of several overhauls of trainee clinical work regulations^18^, their effectiveness in improving patient and resident outcomes has been mixed and heterogenous across training programs.^19^ Moreover, the clinical environment has shifted considerably since the early 2000s. Contemporary trainee work is increasingly dominated by electronic health record (EHR) activity^20^ and digital communication.^21,22^ These tools, while essential, can increase cognitive load and fragmentation of work^23,24^ in ways not reflected in duty hours or simple census count. Concurrently, qualitative and contextual features, such as psychological safety, autonomy, task-switching, rounding structure, clinical acuity, and workspace design, modulate workload but are rarely captured in operational data.^25^ Together, these changes highlight the need for a multidimensional understanding of optimal trainee workload and educational experience, extending beyond single, coarse indicators.

Taking advantage of this dynamic clinical environment, new approaches to understanding the trainee work environment are increasingly available to organizational leaders – including EHR use measures (i.e., digital clickstream data collected during regular work)^20,26–29^ and advanced predictive analytics.^12,30,31^ These emerging tools when paired with brief intermittent surveys offer a pragmatic strategy to link how work is experienced with how it is performed, enabling more precise evaluation of workload, workflow, and educational structures. This approach offers opportunities to provide early signals of strain at the individual or team level, supporting timely intervention.^12^ As such, we conducted a pilot feasibility study to evaluate whether pairing surveys with EHR use data could generate multidimensional, trainee-relevant workload measures that are pragmatic to collect and analytically informative in a real-world graduate medical education setting. This work aimed to lay the groundwork for shaping evidence-based work redesigns that optimize the training experience by maximizing educational value while reducing unnecessary cognitive and administrative burden.

## Methods

### Pilot Design

We performed a mixed-methods pilot to characterize and quantify daily workload and work environment among internal medicine (IM) trainees. The pilot combined daily and weekly survey data and EHR use measures. The pilot was deemed exempt by the Colorado Multiple Institutional Review Board (COMIRB #24-0303). We adhered to the STROBE guidelines for study reporting.^32^

### Setting, participants, and study size

The pilot was conducted between March 31, 2025 and June 22, 2025, at a quaternary care academic hospital with over 700 beds and included IM and medicine-pediatrics residents. Residents were recruited across post-graduate years (PGY) 1-4 and invited to participate by a residency-wide email from a project team member in a non-supervisory role. Eligible residents were those working inpatient services on day shifts that spanned from 6 a.m. to 5 p.m. for seven-day stretches with one day off. Preliminary PGY-1 residents were not eligible for participation. Verbal consent to complete surveys and collect linked anonymized EHR use data was obtained prior to participation. Residents were able to end their participation at any time. Participants received $25 gift cards as compensation for participation in the study after completion of the final survey.

### Survey development

We used the Chokshi and Mann process model^33^ for user-centered digital development to develop and pilot a modified version of a previously described workload survey application^27^ that includes patient census, task load, and health systems culture and validated measurement tools including the National Aeronautics and Space Administration Task Load Index (NASA-TLX)^34^ and the teamwork composite of the Agency of Healthcare Research and Quality (AHRQ) Hospital Survey on Patient Safety Culture (Figure 1).^35^ In phase one of the *discover and define stage*, we identified unique measures of workload that captured a multidimensional view of trainee work using focus groups and a modified Delphi panel.^25^ In phase two, using the previous survey application^27^ as a prototype for this work, we used a mixed methods approach to determine which measures to include for trainees. Specific measures felt to be less relevant to trainees were excluded, and additional measures were incorporated to address previously identified trainee-specific domains of workload, including autonomy, psychological safety, and rounding.^25^ For these domains, validated measurement tools were used when available, including the National Institute for Occupational Safety and Health (NIOSH) Worker Well-Being Questionnaire (WellBQ) on Physical Work Environment Satisfaction,^36^ Professional Well-being Academic Consortium (PWAC) Stanford Professional Fulfillment Index (PFI),^37^ Edmondson Psychological Safety scale,^38^ Work-life climate,^39^ and Moral Distress Thermometer.^40^ Where validated instruments were not available, survey questions were developed and, in phase one of the develop and deliver stage, user-tested with trainees for clarity and consistency. We additionally included a free text question to allow participants to share any other shift satisfiers, dissatisfiers, and notable events. The survey was user-tested among six trainees between January 6, 2025 and January 22, 2025 using structured interviews before finalization (Supplementary Material 1).

In phase two of the develop and deliver stage, we piloted the survey (Supplementary Material 2). Surveys were distributed via REDCap^41^ by email or text message, based on participant preference, at the end of each workday for a total of five daily surveys and one final end-of-week survey. For daily surveys, one reminder was sent after 1.5 hours if no response was received, and two reminders were sent for the end-of-week survey, at 1.5 hours and 24 hours if no response was received.

### EHR data

We extracted aggregated vendor-derived EHR use data (i.e., data collected from regular EHR use during clinical work), including total time in the EHR, secure messaging volume (i.e., number of messages sent and received), and time spent performing various EHR activities, including chart review, documentation, in-basket activities, orders, and problem list management during the pilot week for each participant using Epic Signal.^28,29,42,43^ This data is aggregated at the week-level by participant. Additional more granular EHR use data, including notes signed and first and last login times, was also extracted using pre-existing datasets from the EHR vendor.

### Analysis

Quantitative analyses were performed using SAS Enterprise Guide 9.4 (SAS Institute Inc.). Given the pilot nature of the study and the modest sample size, analyses were limited to unadjusted comparisons and correlations to characterize feasibility and directional associations rather than causal inference. Associations were organized into three domains: (1) subjective shift rating and discrete workload features; (2) correlations between objective workload (e.g., census, NASA-TLX) and subjective domains (environment, professional fulfillment, psychological safety, autonomy, and rounding experience); and (3) concordance between subjective workload and EHR-derived activity metrics (i.e., EHR-derived and granular EHR use data). Associations between the binary shift-rating measure and discrete variables, including patients on treatment team, patients personally responsible for, number of hospital units covered, NASA-TLX scores, and rounding scores, were evaluated using Students’ t-tests.

We then examined correlations among shift-level workload measures. Pearson correlation coefficients were used to assess associations between NASA-TLX scores and, (1) patients on treatment team, (2) patients personally responsible for, and (3) rounding scores.

We further evaluated correlations between each survey-based domain, including WellBQ score, professional fulfillment, work exhaustion, disengagement, psychological safety, work life climate, and moral distress, and shift-level: (1) patients on treatment team, (2) patients personally responsible for, and (3) NASA-TLX.

Finally, we assessed concordance between subjective workload reports and EHR-derived metrics by correlating, (1) self-reported shift hours with total time in the EHR, and (2) reported team and personal patient counts with EHR-derived patient counts. Given the number of comparisons, statistical significance was defined as p<0.002.

To capture subjective components of workload experience not explicitly queried through the survey questionnaire, we completed a hybrid inductive-deductive content analysis of additional free-text comments provided by participants. Four study team members (CF, JC, KR, NB) jointly reviewed comments to develop a codebook (Supplemental Table S2) with working definitions of overarching domains, then individually and separately coded each comment with discrepancies adjudicated as a group to reach consensus.

## Results

### Participants

A total of 37 trainees were eligible and invited for participation in the project, and 28 (76%) trainees participated in the pilot. Trainees included nine (32%) PGY-1, nine (32%) PGY-2, eight (29%) PGY-3, and one (4%) PGY-4. The participants included 14 (50%) trainees in the categorical track, six (21%) in the hospitalist track, five (18%) in the primary care track, and one (4%) in the physician scientist track. Post-graduate year data was missing for one participant, and training track data were missing for two participants. Participant demographics can be found in Table 1.

**Table 1.** Participant demographics.

| **Demographic** | **Participants**<br>**(N = 28, %)** |
| --- | --- |
| **Degree** |  |
| MD | 27 (96) |
| Missing | 1 (4) |
| **Gender** |  |
| Woman | 16 (57) |
| Man | 11 (39) |
| Missing | 1 (4) |
| **Race/Ethnicity<sup>a</sup>** |  |
| American Indian or Alaska Native (e.g., Navajo Nation, Blackfeet Tribe, Inupiat Traditional Gov't., etc.) | 0 (0) |
| Asian or Asian American (e.g., Chinese, Japanese, Filipino, Korean, South Asian, Vietnamese, etc.) | 6 (21) |
| Black or African American (e.g., Jamaican, Nigerian, Haitian, Ethiopian, etc.) | 1 (4) |
| Hispanic or Latino/a (e.g., Puerto Rican, Mexican, Cuban, Salvadoran, Colombian, etc.) | 2 (7) |
| Middle Eastern or North African (e.g., Lebanese, Iranian, Egyptian, Moroccan, Israeli, Palestinian, etc.) | 1 (4) |
| Native Hawai'ian or Pacific Islander (e.g., Samoan, Guamanian, Chamorro, Tongan, etc.) | 0 (0) |
| White or European (e.g., German, Irish, English, Italian, Polish, French, etc.) | 20 (71) |
| Other | 0 (0) |
| Prefer not to answer | 0 (0) |
| Missing | 1 (4) |
| **Post-Graduate Year (PGY)** |  |
| PGY-1 | 9 (32) |
| PGY-2 | 9 (32) |
| PGY-3 | 8 (29) |
| PGY-4 | 1 (4) |
| Missing | 1 (4) |
| **Training track** |  |
| Categorical | 14 (50) |
| Hospitalist | 6 (21) |
| Primary Care | 5 (18) |
| Physician Scientist | 1 (4) |
| Preliminary Year | 0 (0) |
| Missing | 2 (7) |

<sup>a</sup>Multiple selections per participant allowed

### Survey and EHR use data results

Survey responses were completed for all six shifts in the pilot weeks for 26 (93%) participants, with one (4%) participant submitting a survey for four out of the six shifts, and one (4%) participant who completed surveys for five shifts and a partial survey for one shift. A total of 166 shifts were captured. Participants spent 4.4 ± 1.6 (mean ± standard deviation [SD]) minutes completing daily surveys and 8.6 ± 2.3 minutes completing the weekly survey (Supplemental Table S1).

Full survey results can be found in Table 2. Trainees worked a mean of 11.6 ± 1.0 hours per shift and cared for a median of 9 patients (interquartile range [IQR] 6–11) across a median of 3.2 ± 1.1 hospital units. Participants commonly supervised other learners (mean 1.7 ± 1.0 per shift) and most frequently worked with attending physicians (93%), medical students (67%), interns (35%), and senior residents (31%). Hallway rounds were the predominant rounding structure (70%), with bedside rounds reported in 12% of shifts. Mean ratings of rounds on a 1-10 scale reflected moderate educational value (6.1 ± 1.7), length (6.3 ± 1.5), and efficiency (5.7 ± 1.9). Weekly survey results showed low professional fulfillment on average (mean 2.3 ± 0.6), with 11% of participants meeting criteria for fulfillment (score ≥3.0). In contrast, 75% met criteria for work exhaustion and 57% for overall burnout (scores ≥1.33). Psychological safety scores on a 1-7 scale were high (mean 5.4 ± 0.8), and perceived autonomy during shifts closely matched desired autonomy levels. Qualitative content analysis of free-response questions indicated team structure, care coordination, and disruptions/task switching as common contributors to workload (Supplemental Table S2). The remaining free responses can be found in Supplemental Material 3.

**Table 2.**
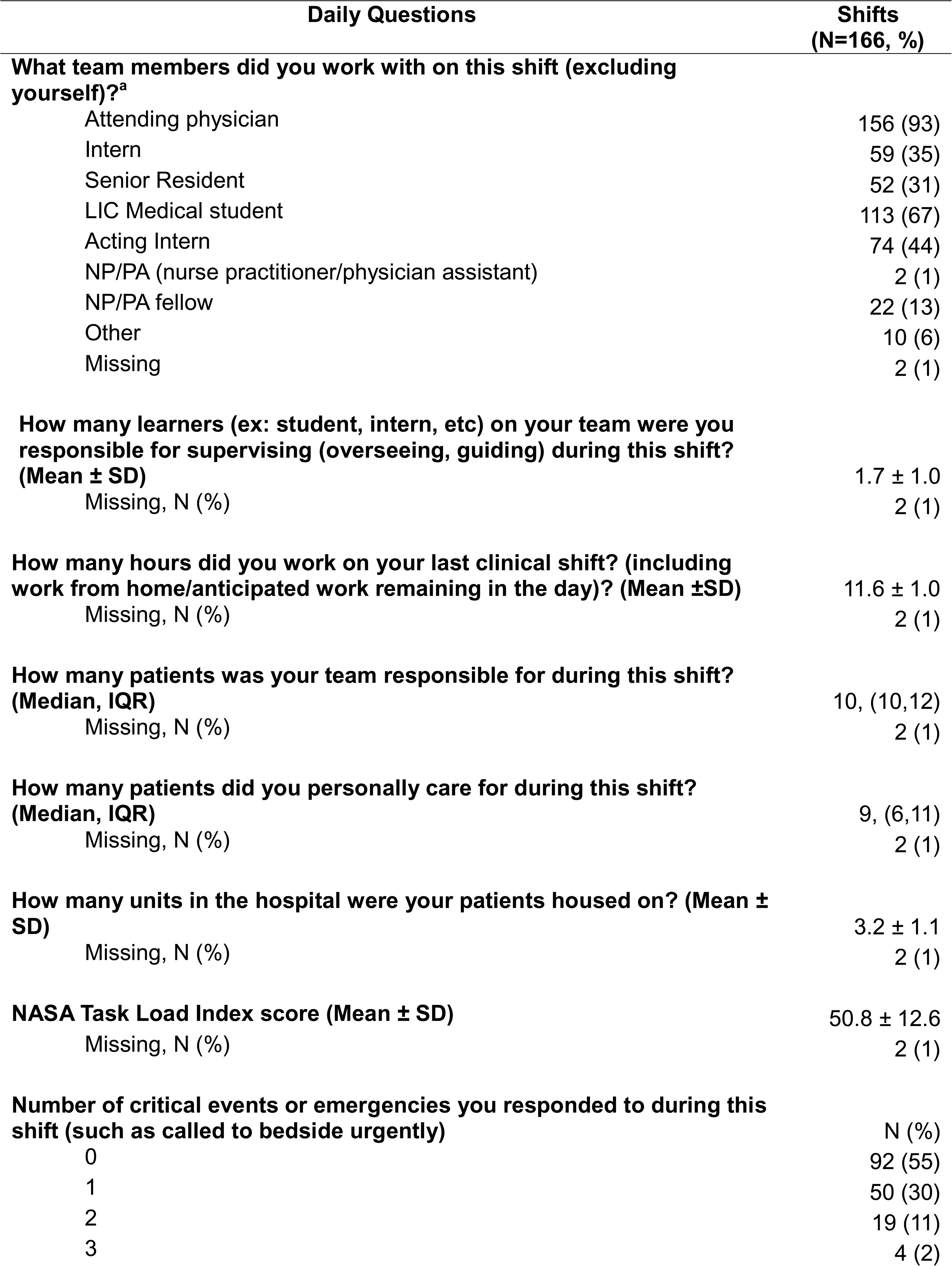
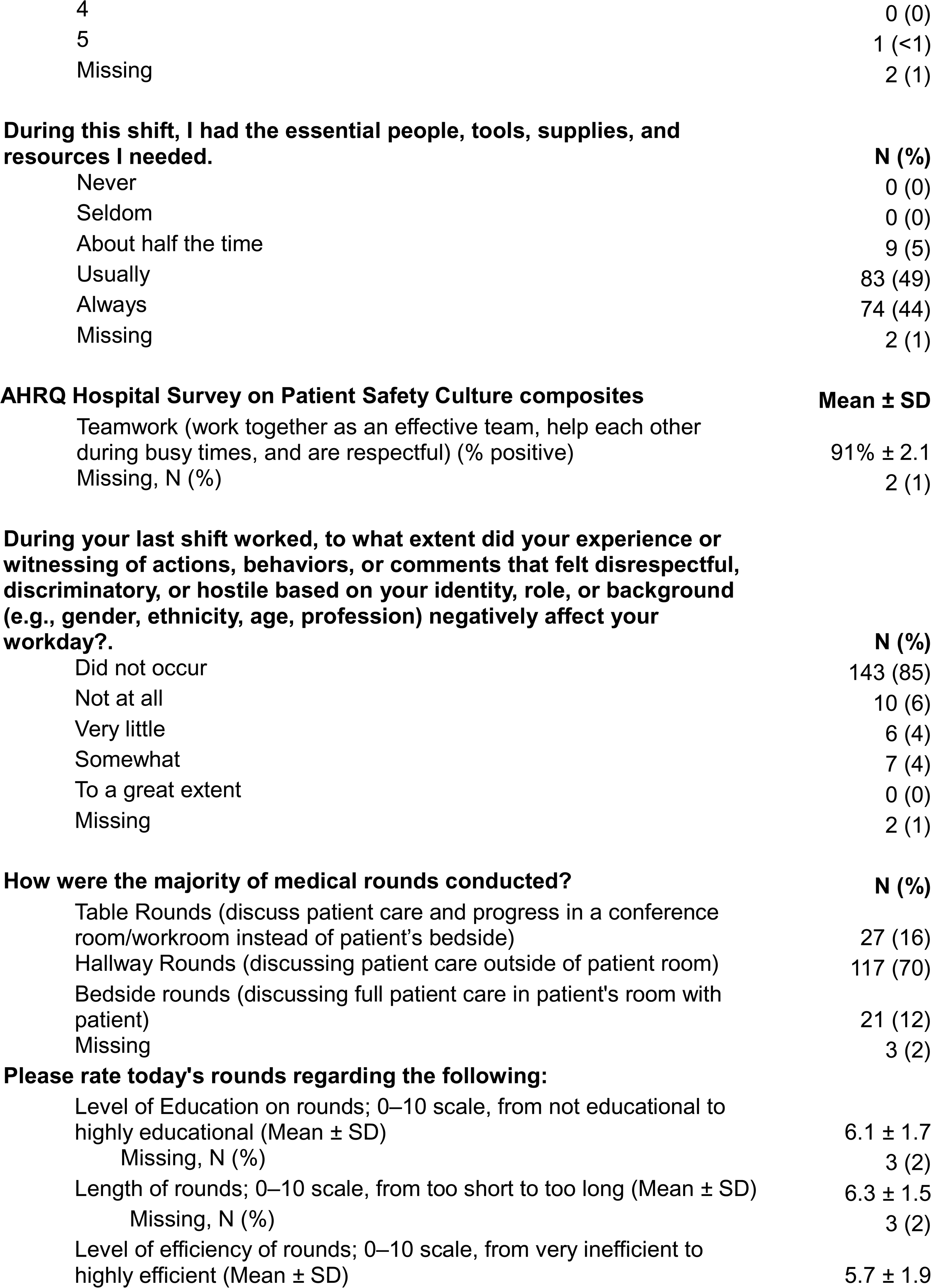
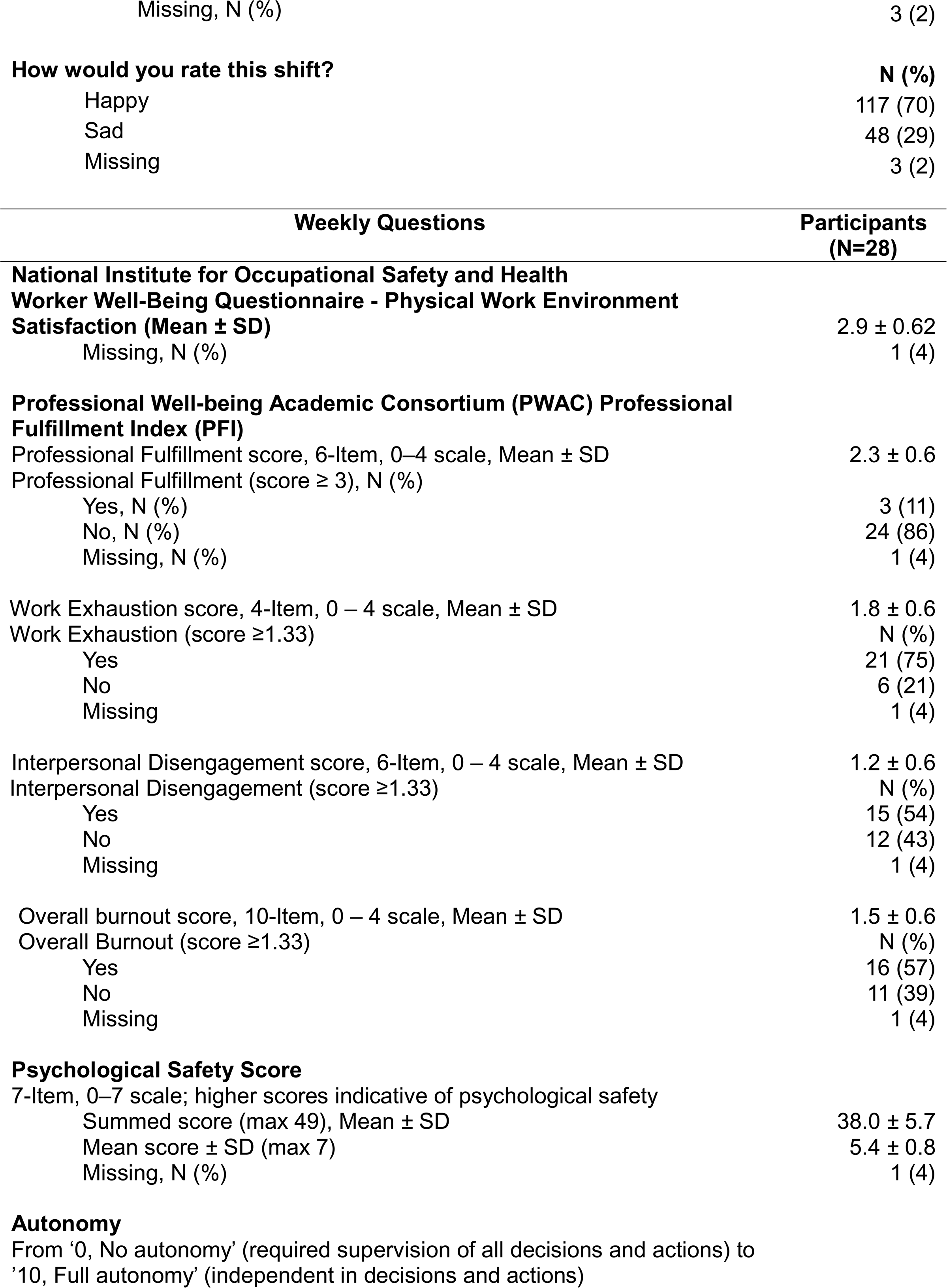
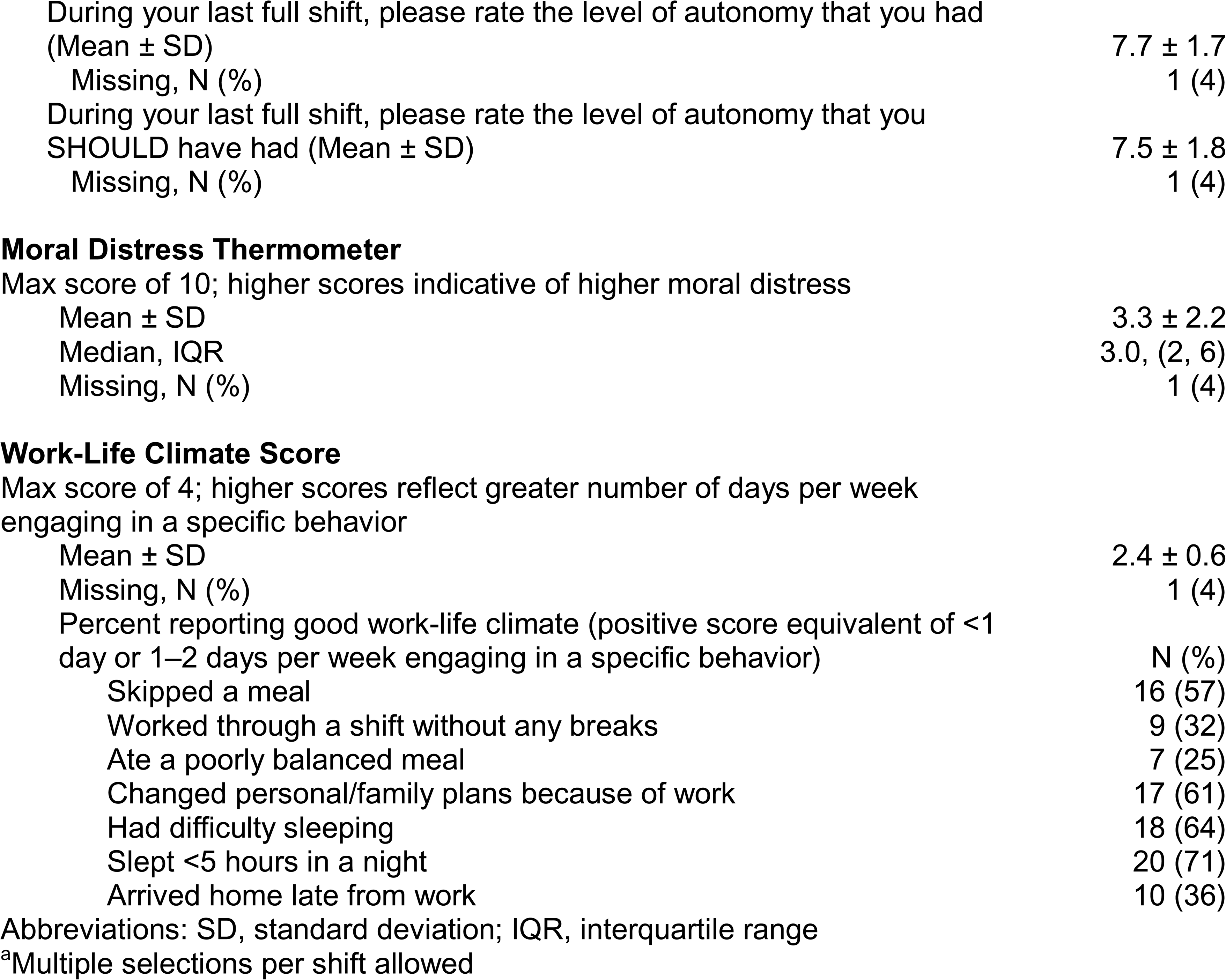
Survey response data.

| Daily Questions | Shifts<br>(N=166, %) |
| --- | --- |
| **What team members did you work with on this shift (excluding yourself)?<sup>a</sup>** |  |
| Attending physician | 156 (93) |
| Intern | 59 (35) |
| Senior Resident | 52 (31) |
| LIC Medical student | 113 (67) |
| Acting Intern | 74 (44) |
| NP/PA (nurse practitioner/physician assistant) | 2 (1) |
| NP/PA fellow | 22 (13) |
| Other | 10 (6) |
| Missing | 2 (1) |
| **How many learners (ex: student, intern, etc) on your team were you responsible for supervising (overseeing, guiding) during this shift? (Mean ± SD)** | 1.7 ± 1.0 |
| Missing, N (%) | 2 (1) |
| **How many hours did you work on your last clinical shift? (including work from home/anticipated work remaining in the day)? (Mean ±SD)** | 11.6 ± 1.0 |
| Missing, N (%) | 2 (1) |
| **How many patients was your team responsible for during this shift? (Median, IQR)** | 10, (10,12) |
| Missing, N (%) | 2 (1) |
| **How many patients did you personally care for during this shift? (Median, IQR)** | 9, (6,11) |
| Missing, N (%) | 2 (1) |
| **How many units in the hospital were your patients housed on? (Mean ± SD)** | 3.2 ± 1.1 |
| Missing, N (%) | 2 (1) |
| **NASA Task Load Index score (Mean ± SD)** | 50.8 ± 12.6 |
| Missing, N (%) | 2 (1) |
| **Number of critical events or emergencies you responded to during this shift (such as called to bedside urgently)** | N (%) |
| 0 | 92 (55) |
| 1 | 50 (30) |
| 2 | 19 (11) |
| 3 | 4 (2) |
| 4 | 0 (0) |
| 5 | 1 (<1) |
| Missing | 2 (1) |

**During this shift, I had the essential people, tools, supplies, and resources I needed.**
|  | **N (%)** |
| --- | --- |
| Never | 0 (0) |
| Seldom | 0 (0) |
| About half the time | 9 (5) |
| Usually | 83 (49) |
| Always | 74 (44) |
| Missing | 2 (1) |
| **Mean ± SD** |  |
| Teamwork (work together as an effective team, help each other during busy times, and are respectful) (% positive) | 91% ± 2.1 |
| Missing, N (%) | 2 (1) |

|  | **N (%)** |
| --- | --- |
| Did not occur | 143 (85) |
| Not at all | 10 (6) |
| Very little | 6 (4) |
| Somewhat | 7 (4) |
| To a great extent | 0 (0) |
| Missing | 2 (1) |
| Table Rounds (discuss patient care and progress in a conference room/workroom instead of patient's bedside) | 27 (16) |
| Hallway Rounds (discussing patient care outside of patient room) | 117 (70) |
| Bedside rounds (discussing full patient care in patient's room with patient) | 21 (12) |
| Missing | 3 (2) |
| **Please rate today's rounds regarding the following:** |  |
| Level of Education on rounds; 0–10 scale, from not educational to highly educational (Mean ± SD) | 6.1 ± 1.7 |
| Missing, N (%) | 3 (2) |
| Length of rounds; 0–10 scale, from too short to too long (Mean ± SD) | 6.3 ± 1.5 |
| Missing, N (%) | 3 (2) |
| Level of efficiency of rounds; 0–10 scale, from very inefficient to highly efficient (Mean ± SD) | 5.7 ± 1.9 |
| Missing, N (%) | 3 (2) |
| **How would you rate this shift?** | **N (%)** |
| Happy | 117 (70) |
| Sad | 48 (29) |
| Missing | 3 (2) |

| Weekly Questions | Participants<br>(N=28) |
| --- | --- |
| **National Institute for Occupational Safety and Health**<br>**Worker Well-Being Questionnaire - Physical Work Environment**<br>**Satisfaction (Mean ± SD)** | 2.9 ± 0.62 |
| Missing, N (%) | 1 (4) |
| **Professional Well-being Academic Consortium (PWAC) Professional**<br>**Fulfillment Index (PFI)** |  |
| Professional Fulfillment score, 6-Item, 0–4 scale, Mean ± SD | 2.3 ± 0.6 |
| Professional Fulfillment (score ≥ 3), N (%) | |
| Yes, N (%) | 3 (11) |
| No, N (%) | 24 (86) |
| Missing, N (%) | 1 (4) |
| Work Exhaustion score, 4-Item, 0 – 4 scale, Mean ± SD | 1.8 ± 0.6 |
| Work Exhaustion (score ≥1.33) | N (%) |
| Yes | 21 (75) |
| No | 6 (21) |
| Missing | 1 (4) |
| Interpersonal Disengagement score, 6-Item, 0 – 4 scale, Mean ± SD | 1.2 ± 0.6 |
| Interpersonal Disengagement (score ≥1.33) | N (%) |
| Yes | 15 (54) |
| No | 12 (43) |
| Missing | 1 (4) |
| Overall burnout score, 10-Item, 0 – 4 scale, Mean ± SD | 1.5 ± 0.6 |
| Overall Burnout (score ≥1.33) | N (%) |
| Yes | 16 (57) |
| No | 11 (39) |
| Missing | 1 (4) |
| **Psychological Safety Score** |  |
| 7-Item, 0–7 scale; higher scores indicative of psychological safety |  |
| Summed score (max 49), Mean ± SD | 38.0 ± 5.7 |
| Mean score ± SD (max 7) | 5.4 ± 0.8 |
| Missing, N (%) | 1 (4) |
| During your last full shift, please rate the level of autonomy that you had (Mean ± SD) | 7.7 ± 1.7 |
| Missing, N (%) | 1 (4) |
| During your last full shift, please rate the level of autonomy that you SHOULD have had (Mean ± SD) | 7.5 ± 1.8 |
| Missing, N (%) | 1 (4) |
| Mean ± SD | 3.3 ± 2.2 |
| Median, IQR | 3.0, (2, 6) |
| Missing, N (%) | 1 (4) |
| Mean ± SD | 2.4 ± 0.6 |
| Missing, N (%) | 1 (4) |
| Percent reporting good work-life climate (positive score equivalent of <1 day or 1–2 days per week engaging in a specific behavior) | N (%) |
| Skipped a meal | 16 (57) |
| Worked through a shift without any breaks | 9 (32) |
| Ate a poorly balanced meal | 7 (25) |
| Changed personal/family plans because of work | 17 (61) |
| Had difficulty sleeping | 18 (64) |
| Slept <5 hours in a night | 20 (71) |
| Arrived home late from work | 10 (36) |

Abbreviations: SD, standard deviation; IQR, interquartile range
<sup>a</sup>Multiple selections per shift allowed

We found that the mean NASA-TLX score was lower among shifts rated with “happy face” emojis compared to those with “sad face” emojis (46 ± 11 vs. 62 ± 9; p<0.0001) (Supplemental Table S3, Supplemental Figure S1). Similarly, mean rounding length scores were lower among shifts rated with “happy face” emojis compared to those with “sad face” emojis (6 ± 1 vs. 7 ± 2, p<0.0001) (Supplemental Table S4, Supplemental Figure S2). Mean rounding length scores were correlated with mean NASA-TLX scores (0.35 [95% CI: 0.21, 0.48], p<0.0001) (Supplemental Figure S3). No significant associations were found when comparing number of patients on the treatment team/patients personally responsible with shift rating, NASA-TLX, WellBQ score, professional fulfillment, work exhaustion, disengagement, psychological safety, work life climate, or moral distress scores. Similarly, NASA-TLX was not associated with any of these survey-based domains.

In regard to EHR use data, time from initial login to last login among trainees was 15 ± 2 hours per day (Table 3). Trainees spent an average of 204 ± 46 minutes per day in the EHR, primarily spent on documentation, chart review, and order management. Trainees signed 6 ± 3 notes per day. We found no associations between reported shift hours, and total time spent in the EHR but did find a correlation between reported shift hours and duration of shift measured as time from initial to last login into the EHR (*r*=0.43 [95% CI: 0.30, 0.55], p<0.0001). We also found a correlation between mean number of notes signed per day and mean reported team census (*r*=0.19 [95% CI: 0.04, 0.33], p=0.0130) and personal census (*r*=0.34 [95% CI: 0.20, 0.47], p<0.0001). Similarly, we found no associations between EHR-based identification of number of patients cared for, which was defined as average count of patients per day where the provider has co-signed, attested, or contributed characters to a note, and survey reports of number of patients on treatment team or personally responsible for.

**Table 3.** EHR use data.

| **Metric** | **Mean ± SD** |
| --- | --- |
| Patients per Day Value<sup>a</sup> | 5 ± 1 |
| Patients per Day Denominator<sup>b</sup> | 7 ± 1 |
| Notes per day<sup>c</sup> | 6 ± 3 |
| Duration of shift<sup>d</sup> | 15 ± 2 |
| **Messages per day (Mean ± SD)** |  |
| Secure Messages Sent | 53 ± 19 |
| Secure Messages Received | 111 ± 34 |
| **Time (mins) per day (Mean ± SD)** |  |
| All time provider was logged into the system<sup>e</sup> | 204 ± 46 |
| Chart Review | 13 ± 13 |
| Clinical Review | 55 ± 17 |
| Documentation | 57 ± 19 |
| In Basket | 0.5 ± 0.4 |
| Notes | 57 ± 19 |
| Orders | 33 ± 9 |
| Problem List | 0.3 ± 0.2 |

Abbreviations: SD, standard deviation
<sup>a</sup>Average count of patients per day where the provider has co-signed, attested, or contributed characters to a note.
<sup>b</sup>Sum of patients (for whom the provider co-signed, attested, or contributed at least one character to a note) over each day within the reporting period.
<sup>c</sup>Average count of notes signed per shift.
<sup>d</sup>Duration in hours between first and last electronic health record log in time per shift
<sup>e</sup>Includes mobile time

## Discussion

In this pilot feasibility study, we sought to determine whether regular, brief trainee surveys could be paired with EHR use data to capture multidimensional aspects of trainee workload in a pragmatic manner. We found high engagement with the survey instrument, minimal time burden, and positive feedback regarding continued participation, supporting feasibility in routine clinical practice. Subjective measures, including shift ratings and NASA-TLX scores, demonstrated consistent relationships with rounding duration, suggesting sensitivity to variations in cognitive load. EHR use data provided complementary, objective measures of task-level work that contextualized these insights. Together, these findings indicate that paired subjective and objective workload measurement is feasible in graduate medical education and yields information not captured by traditional metrics alone. These complementary data streams have the potential to lead to timely and meaningful change, in contrast to the prolonged runway to await policy or regulatory changes that have traditionally informed the structure of trainee work.

Although this pilot was not powered to test causal associations, several descriptive patterns aligned with existing literature and warrant consideration. Rounding emerged as a prominent contributor to perceived workload, with subjectively longer rounding duration associated with poorer shift ratings and higher cognitive load. Trainees frequently described prolonged and inefficient rounding practices with limited educational value, consistent with prior studies demonstrating resident rounding has substantially lengthened in recent years.^44–46^ Notably, in comparison to recent evidence among hospitalists,^27^ cognitive load among trainees was generally higher. These findings highlight rounding structure and cognitive load as potentially high-yield targets for future work design interventions.

EHR use data further demonstrated substantial time devoted to documentation, orders, chart review, and secure messaging volume, activities that consume cognitive bandwidth but are largely invisible to traditional conventional workload metrics. Both secure messaging and disruptions/task switching, which are known to be associated, emerged as workload domains in qualitative content analysis of free-text survey responses, and align with prior work suggesting that secure messaging volume is associated with EHR wrong-patient ordering errors^47^ and nonurgent and non-actionable secure messages are intervenable targets for improvement.^22^ Moreover, residents spent significantly more daily time in the EHR compared to hospitalists^27^, underscoring both the administrative burden borne by trainees and the importance of objective measures that capture digital work. Total EHR time in our study was significantly less than survey-based reports of total time spent at work, similar to previous findings.^27^ EHR-derived measures capture observable EHR interactions but may under-represent cognitive work performed outside of the system and can vary by vendor algorithms for “time-outs” due to pauses in mouse/keystroke data.^26^ This may explain the identified correlation between reported hours and time in EHR measured between first and last log-in, which avoids “time-out” issues, but may overestimate continuous work if sessions are interrupted (e.g. off-site logins). Together, these findings suggest that not all EHR-derived metrics reflect lived workload equally. Measures that capture the span of work (i.e., first-to-last login) aligned more closely with trainee-reported hours than aggregated active EHR time, highlighting the importance of selecting workload measures that map to how work is experienced.

The operational implications of this feasibility study suggest that integrated dashboards that combine survey-based signals with EHR-derived indicators could support more responsive approaches to monitoring trainee workload. When paired with brief, intermittent surveys that capture contextual features of work, these complementary data streams may enable earlier identification of emerging strain for teams and for specific rotations, as well as the empirical derivation of optimal work and educational design, well before downstream outcomes such as burnout or attrition occur. Beyond surveillance, these data can serve as baseline metrics to evaluate targeted interventions, such as restructuring rounds, optimizing messaging workflows, or streamlining documentation processes. Importantly, implementation of monitoring systems should be coupled with clear, non-punitive escalation pathways to ensure that identified workload concerns prompt timely support rather than unintended consequences.

### Limitations

This study has several limitations to consider. First, our study population was limited to a single residency program with a small sample size and may reflect institution-specific findings rather than generalizable trends. Our study was conducted among IM and medicine-pediatrics residents assigned to inpatient ward primary services during the daytime and may not represent non-IM contexts nor be generalizable to outpatient settings or nocturnal coverage. Although survey data may suffer from response bias, our study sought to additionally evaluate EHR data as a measure of workload. Finally, given our study was conducted by an opt-in survey, it may suffer from participation bias. Our survey findings should be interpreted cautiously and considered hypothesis-generating and merit more rigorous testing through larger, multicenter studies and implementation trials in varying contexts.

## Conclusion

Combining brief surveys with granular EHR audit-log data provides a feasible, practical, and scalable approach to capturing multidimensional aspects of trainee workload in real-world clinical settings. This paired measurement strategy extends beyond traditional indicators by integrating subjective experience with objective task-level work, offering actionable insight into how workload is structured and experienced. In this pilot, these data highlighted potentially modifiable contributors to workload, including cognitive load, rounding structure, and EHR-related burden, and demonstrated acceptability and analytic utility in graduate medical education. Embedding such measurement into routine practice may provide a foundation for evidence-based work design efforts aimed at improving the clinical learning environment while reducing unnecessary cognitive and administrative strain. Future studies should consider leveraging these tools to power trials of targeted work redesign and to evaluate downstream effects on education, clinician well-being, and patient outcomes.

## Acknowledgements and Conflicts of Interest

Drs. Burden and Keniston report funding from the Agency for Healthcare Research and Quality, the National Institute for Occupational Health and Safety, University of Colorado Innovations digiSPARK award, and the American Medical Association not related to this work.

Drs. Burden and Keniston contributed to the development of GrittyWork, a digital workforce application, and a registered trademark of the University of Colorado. Dr. Burden also reports funding from Med-IQ and CDC NIOSH Center for Health, Work, and Environment (CHWE), a NIOSH Center of Excellence for Total Worker Health not related to this work. Dr. Burden reports funding from CU Thrive Precision Innovation Award.

The authors utilized the ChatGPT language model developed by OpenAI and Microsoft Copilot for editing original author content to improve readability. All information and materials in the manuscript are original.

Dr. Karan Rai had access to all the data in the project and takes responsibility for the integrity of the data and the accuracy of the data analysis.

The views expressed in this article are those of the authors and do not necessarily represent the views, policies, or positions of any affiliated institutions or employers. Similarly, concepts presented in the introduction and discussion are intended to illustrate general principles and do not describe or reflect any specific health system or organization.

## Ethical approval

This study was deemed exempt by the Colorado Multiple Institutional Review Board (COMIRB#24-0303)

## Previous presentations

none

## Data availability

The data underlying this article will be shared on reasonable request to the corresponding author.

---

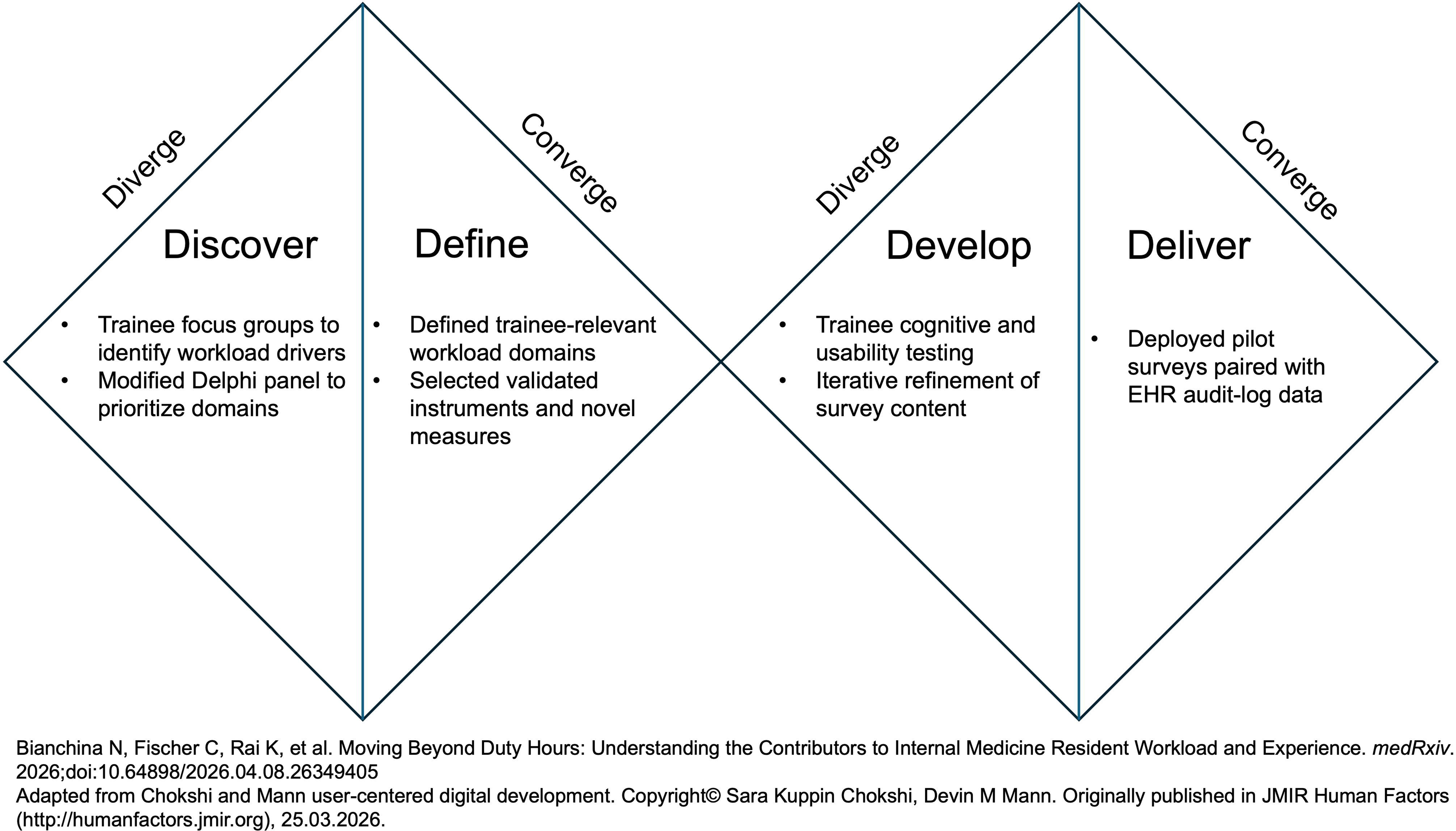

## References

1. Elliott DJ, Young RS, Brice J, Aguiar R, Kolm P. Effect of hospitalist workload on the quality and efficiency of care. JAMA Intern Med. May 2014;174(5):786–93. doi:10.1001/jamainternmed.2014.300

2. Sevransky JE, Chai ZJ, Cotsonis GA, Cobb JP, Pastores SM. Survey of Annual Staffing Workloads for Adult Critical Care Physicians Working in the United States. Ann Am Thorac Soc. May 2016;13(5):751–3. doi:10.1513/AnnalsATS.201508-502LE

3. West CP, Dyrbye LN, Shanafelt TD. Physician burnout: contributors, consequences and solutions. J Intern Med. Jun 2018;283(6):516–529. doi:10.1111/joim.12752

4. Fang Y, Lodi S, Hughes TM, Frank E, Sen S, Bohnert ASB. Work Hours and Depression in U.S. First-Year Physicians. N Engl J Med. Oct 20 2022;387(16):1522–1524. doi:10.1056/NEJMc2210365

5. van der Heijden F, Dillingh G, Bakker A, Prins J. Suicidal thoughts among medical residents with burnout. Arch Suicide Res. 2008;12(4):344–6. doi:10.1080/13811110802325349

6. Shanafelt T, Goh J, Sinsky C. The Business Case for Investing in Physician Well-being. JAMA Intern Med. Dec 1 2017;177(12):1826–1832. doi:10.1001/jamainternmed.2017.4340

7. Ahmed A, Podolsky SH. House Staff Unionization - A Historical Tool Revisited. N Engl J Med. Sep 19 2024;391(11):1060–1064. doi:10.1056/NEJMms2404695

8. Rosenbaum L. What Do Trainees Want? The Rise of House Staff Unions. N Engl J Med. Jan 18 2024;390(3):279–283. doi:10.1056/NEJMms2308224

9. Lin GL, Ge TJ, Pal R. Resident and Fellow Unions: Collective Activism to Promote Well-being for Physicians in Training. JAMA. Aug 16 2022;328(7):619–620. doi:10.1001/jama.2022.12838

10. Brajcich BC, Chung JW, Wood DE, et al. National Evaluation of the Association Between Resident Labor Union Participation and Surgical Resident Well-being. JAMA Netw Open. Sep 1 2021;4(9):e2123412. doi:10.1001/jamanetworkopen.2021.23412

11. Foote DC, Rosenblatt AE, Amortegui D, et al. Experiences With Unionization Among General Surgery Resident Physicians, Faculty, and Staff. JAMA Netw Open. Jul 1 2024;7(7):e2421676. doi:10.1001/jamanetworkopen.2024.21676

12. Burden M, Dyrbye L. Evidence-Based Work Design - Bridging the Divide. N Engl J Med. Mar 13 2025;392(11):1044–1046. doi:10.1056/NEJMp2412389

13. Burden M, Patel M, Kissler M, Harry E, Keniston A. Measuring and driving hospitalist value: Expanding beyond wRVUs. J Hosp Med. Sep 2022;17(9):760–764. doi:10.1002/jhm.12849

14. Kulkarni SA, Keniston A, Linker AS, et al. Building a thriving academic hospitalist workforce: A rapid qualitative analysis identifying key areas of focus in the field. J Hosp Med. Apr 2023;18(4):329–336. doi:10.1002/jhm.13074

15. Coit MH, Katz JT, McMahon GT. The effect of workload reduction on the quality of residents’ discharge summaries. J Gen Intern Med. Jan 2011;26(1):28–32. doi:10.1007/s11606-010-1465-z

16. Burden M, McBeth L, Keniston A. Salient Measures of Hospitalist Workload. JAMA Netw Open. Aug 1 2023;6(8):e2328165. doi:10.1001/jamanetworkopen.2023.28165

17. Fletcher KE, Visotcky AM, Slagle JM, et al. Self-reported subjective workload of on-call interns. J Grad Med Educ. Sep 2013;5(3):427–32. doi:10.4300/JGME-D-12-00241.1

18. Spritz N. Oversight of physicians’ conduct by state licensing agencies. Lessons from New York’s Libby Zion case. Ann Intern Med. Aug 1 1991;115(3):219–22. doi:10.7326/0003-4819-115-3-219

19. Lark ME, Airhart AE, Miller PM, Chung JW, Bilimoria KY. Accreditation Council for Graduate Medical Education Work Hour Requirements: A Review of Recent Evidence. J Grad Med Educ. Oct 2025;17(5):676–681. doi:10.4300/JGME-D-25-00784.1

20. Wang JK, Ouyang D, Hom J, Chi J, Chen JH. Characterizing electronic health record usage patterns of inpatient medicine residents using event log data. PLoS One. 2019;14(2):e0205379. doi:10.1371/journal.pone.0205379

21. Brickson C, Keniston A, Knees M, Burden M. Characterizing electronic messaging use among hospitalists and its association with patient volumes. J Hosp Med. Dec 2024;19(12):1131–1137. doi:10.1002/jhm.13462

22. Clawson J, Grobbel E, Keniston A, McBeth L, Knees M, Burden M. Message madness: Characterization of electronic secure messages in the hospital. J Hosp Med. Aug 7 2025;doi:10.1002/jhm.70144

23. Kochar A, Knees M, Arnold RM. Training Interrupted-The Hidden Costs of Secure Messaging on Medical Trainee Learning. JAMA Intern Med. Jul 1 2025;185(7):761–762. doi:10.1001/jamainternmed.2025.1245

24. Knees M, Keniston A, Yu A, et al. Academic hospitalist perspectives on the benefits and challenges of secure messaging: A mixed methods analysis. J Hosp Med. Mar 2025;20(3):248–257. doi:10.1002/jhm.13522

25. Bianchina N, Fischer C, Rai K, et al. Moving Beyond Duty Hours: Understanding the Contributors to Internal Medicine Resident Workload and Experience. medRxiv. 2026;doi:10.64898/2026.04.08.26349405

26. Burden M, Keniston A, Pell J, Yu A, Dyrbye L, Kannampallil T. Unlocking inpatient workload insights with electronic health record event logs. J Hosp Med. Jan 2025;20(1):79–84. doi:10.1002/jhm.13386

27. Burden M, McBeth L, Keniston A. The development and pilot of a novel mobile application to assess clinician perception of workload and work environment. J Hosp Med. Aug 2024;19(8):661–670. doi:10.1002/jhm.13366

28. Lou SS, Liu H, Warner BC, Harford D, Lu C, Kannampallil T. Predicting physician burnout using clinical activity logs: Model performance and lessons learned. J Biomed Inform. Mar 2022;127:104015. doi:10.1016/j.jbi.2022.104015

29. Adler-Milstein J, Adelman JS, Tai-Seale M, Patel VL, Dymek C. EHR audit logs: A new goldmine for health services research? J Biomed Inform. Jan 2020;101:103343. doi:10.1016/j.jbi.2019.103343

30. Baratta LR, Lou SS, Kannampallil T, Jha S, Roy A, Cobb AD. Predicting Secure Messaging Traffic in Clinical Settings. Stud Health Technol Inform. Aug 7 2025;329:553–557. doi:10.3233/SHTI250901

31. Kim S, Lou SS, Baratta LR, Kannampallil T. Classifying clinical work settings using EHR audit logs: a machine learning approach. Am J Manag Care. Jan 1 2023;29(1):e24–e30. doi:10.37765/ajmc.2023.89310

32. von Elm E, Altman DG, Egger M, et al. The Strengthening the Reporting of Observational Studies in Epidemiology (STROBE) statement: guidelines for reporting observational studies. J Clin Epidemiol. Apr 2008;61(4):344–9. doi:10.1016/j.jclinepi.2007.11.008

33. Chokshi SK, Mann DM. Innovating From Within: A Process Model for User-Centered Digital Development in Academic Medical Centers. JMIR Hum Factors. Dec 19 2018;5(4):e11048. doi:10.2196/11048

34. Hart SG, Staveland LE. Development of NASA-TLX (Task Load Index): Results of Empirical and Theoretical Research. Advances in Psychology. 1988;52:139–183. doi:10.1016/S0166-4115(08)62386-9

35. Zebrak K, Yount N, Sorra J, et al. Development, Pilot Study, and Psychometric Analysis of the AHRQ Surveys on Patient Safety Culture (SOPS((R))) Workplace Safety Supplemental Items for Hospitals. Int J Environ Res Public Health. Jun 2 2022;19(11)doi:10.3390/ijerph19116815

36. Chari R CC-C, Sauter SL, Petrun Sayers EL, Huang W, Fisher GG. NIOSH worker well-being questionnaire (WellBQ). In: U.S. Department of Health and Human Services CfDCaP, National Institute for Occupational Safety and Health, DHHS (NIOSH), editor. Cincinnati, OH2021.

37. Trockel M, Bohman B, Lesure E, et al. A Brief Instrument to Assess Both Burnout and Professional Fulfillment in Physicians: Reliability and Validity, Including Correlation with Self-Reported Medical Errors, in a Sample of Resident and Practicing Physicians. Acad Psychiatry. Feb 2018;42(1):11–24. doi:10.1007/s40596-017-0849-3

38. Edmondson A. Psychological Safety and Learning Behavior in Work Teams. Administrative Science Quarterly. 2016;44(2):350–383. doi:10.2307/2666999

39. Sexton JB, Schwartz SP, Chadwick WA, et al. The associations between work-life balance behaviours, teamwork climate and safety climate: cross-sectional survey introducing the work-life climate scale, psychometric properties, benchmarking data and future directions. BMJ Qual Saf. Aug 2017;26(8):632–640. doi:10.1136/bmjqs-2016-006032

40. Wocial LD, Weaver MT. Development and psychometric testing of a new tool for detecting moral distress: the Moral Distress Thermometer. J Adv Nurs. Jan 2013;69(1):167–74. doi:10.1111/j.1365-2648.2012.06036.x

41. Harris PA, Taylor R, Thielke R, Payne J, Gonzalez N, Conde JG. Research electronic data capture (REDCap)--a metadata-driven methodology and workflow process for providing translational research informatics support. J Biomed Inform. Apr 2009;42(2):377–81. doi:10.1016/j.jbi.2008.08.010

42. Cohen GR, Boi J, Johnson C, Brown L, Patel V. Measuring time clinicians spend using EHRs in the inpatient setting: a national, mixed-methods study. J Am Med Inform Assoc. Jul 30 2021;28(8):1676–1682. doi:10.1093/jamia/ocab042

43. Dyrbye LN, Gordon J, O’Horo J, et al. Relationships Between EHR-Based Audit Log Data and Physician Burnout and Clinical Practice Process Measures. Mayo Clin Proc. Mar 2023;98(3):398–409. doi:10.1016/j.mayocp.2022.10.027

44. Stickrath C, Noble M, Prochazka A, et al. Attending rounds in the current era: what is and is not happening. JAMA Intern Med. Jun 24 2013;173(12):1084–9. doi:10.1001/jamainternmed.2013.6041

45. Miller M, Johnson B, Greene HL, Baier M, Nowlin S. An observational study of attending rounds. J Gen Intern Med. Nov-Dec 1992;7(6):646–8. doi:10.1007/BF02599208

46. Lau CJ, C; Apgar, S; Marano, P; Lai, AR; Khan, A. HOW DID YOU SPEND YOUR TIME TODAY? SIMILARITIES AND DIFFERENCES IN CLINICAL WORK OF MEDICINE INTERNS AND DIRECT CARE HOSPITALISTS. presented at: Hospital Medicine 2020; 2020; Virtual. Accessed November 10, 2025.

47. Lou SS, Lew D, Xia L, Baratta L, Eiden E, Kannampallil T. Secure Messaging Use and Wrong-Patient Ordering Errors Among Inpatient Clinicians. JAMA Netw Open. Dec 2 2024;7(12):e2447797. doi:10.1001/jamanetworkopen.2024.47797

