## Supplemental Material for "Characterizing Trainee Workload Using Paired Daily Surveys and EHR Use Data: A Mixed-Methods Pilot Study"

TABLE OF CONTENTS

Supplementary Material 1. Survey user testing interview script

Supplementary Material 2. Full survey

Supplemental Table S1. Post-pilot Survey User Experience Evaluation

Supplemental Table S2. Qualitative content analysis of free text responses

Supplemental Material 3. Survey Free Text Responses

Supplemental Table S3. NASA-TLX scores by shift rating

Supplemental Figure S1. Box plots for NASA-TLX scores by shift rating

Supplemental Table S4. Rounding length score by shift rating

Supplemental Figure S2. Box plots for rounding length score by shift rating

Supplemental Figure S3. Scatter plot for correlation between length of rounds score and NASA-TLX score

Supplementary material 1. Survey user testing interview script

Introduction

Currently there is very little research exploring what contributes to resident workload and how we best measure this. We know that a resident’s job is dramatically different from that of an attending clinician and may need different approaches to measure the workload associated with this role. We are exploring contributors to your workload and ideas or challenges in measuring this. We have conducted initial focus groups evaluating this and created the following survey we are asking you to evaluate. The goal is to have a daily, weekly and monthly survey to understand a trainee workload in real-time with future potential to alter this daily to maximize resident thriving. We are seeking your feedback in our survey questions.

Your participation is strictly voluntary, and all your responses will be completely anonymous. It is important that what is said here remains confidential so that we can create a safe environment to speak freely without judgment or negative repercussions. We will record our discussion today as we want to be able to capture everything that is said, but we will not be reporting on one specific person or disclosing any information that will identify a person, program, or specific location in the transcript or final reports.

We would like to audio-record these groups to summarize. We will disseminate this information amongst our project team and potentially use the information as part of future summaries or presentations. We will NOT use the recordings for any other purpose, send them to anyone else, or quote them by name. You can choose not to participate in the small group discussions, and if you decide to participate, you do not have to speak and can leave at any time.

When you are done reviewing the questions, please raise your hand so we know you are finished. If you do not have any

Does everyone agree to these ground rules? Do you have any questions? Are you ready to get started?

Focus Group/Interview Guide

**Section 1:**

After each question: What is the question trying to find out from you?

After question 1 – do you view this to include WFH?

After all are done: What, if anything, was unclear or confusing about the question or the way you would answer?

**Section 2:**

NASA-TLX: What, if anything, was unclear or confusing? What’s your opinion on how the information is laid out?

Question 7-12:  What is the question trying to find out from you?

After all questions: What if anything was unclear or confusing?

**Section 3:**

Questions 13-14: What is the question trying to find out from you?

After all questions: What if anything was unclear or confusing?

**Section 4:**

Question 15-17: What is the question trying to find out from you? What, if anything, was unclear or confusing about the question or the way you would answer?

Questions 17- 18: What is the question trying to find out from you? How did you come up with your answer?

**Section 5:**

Question 1: What, if anything, was unclear or confusing about the question or the way you would answer?

Question 2: What, if anything, was unclear or confusing about the question or the way you would answer?

Question 3:  What, if anything, was unclear or confusing about the question or the way you would answer?

Question 4: What, if anything, was unclear or confusing about the question or the way you would answer?

**Section 6:**

Psych Safety: What is your first impression of this series of questions? What, if anything, was unclear or confusing about the question or the way you would answer?

Autonomy: Answer the question. How did you get to that answer. What is your first impression of this series of questions? What, if anything, was unclear or confusing about the question or the way you would answer?

**Section 7:**

Question 15: What is the question trying to find out from you?

Question 16: What is the question trying to find out from you?

Question 17: What is the question trying to find out from you?

Question 15-17: What, if anything, was unclear or confusing about the question or the way you would answer?

Supplementary Material 2. Daily and weekly surveys
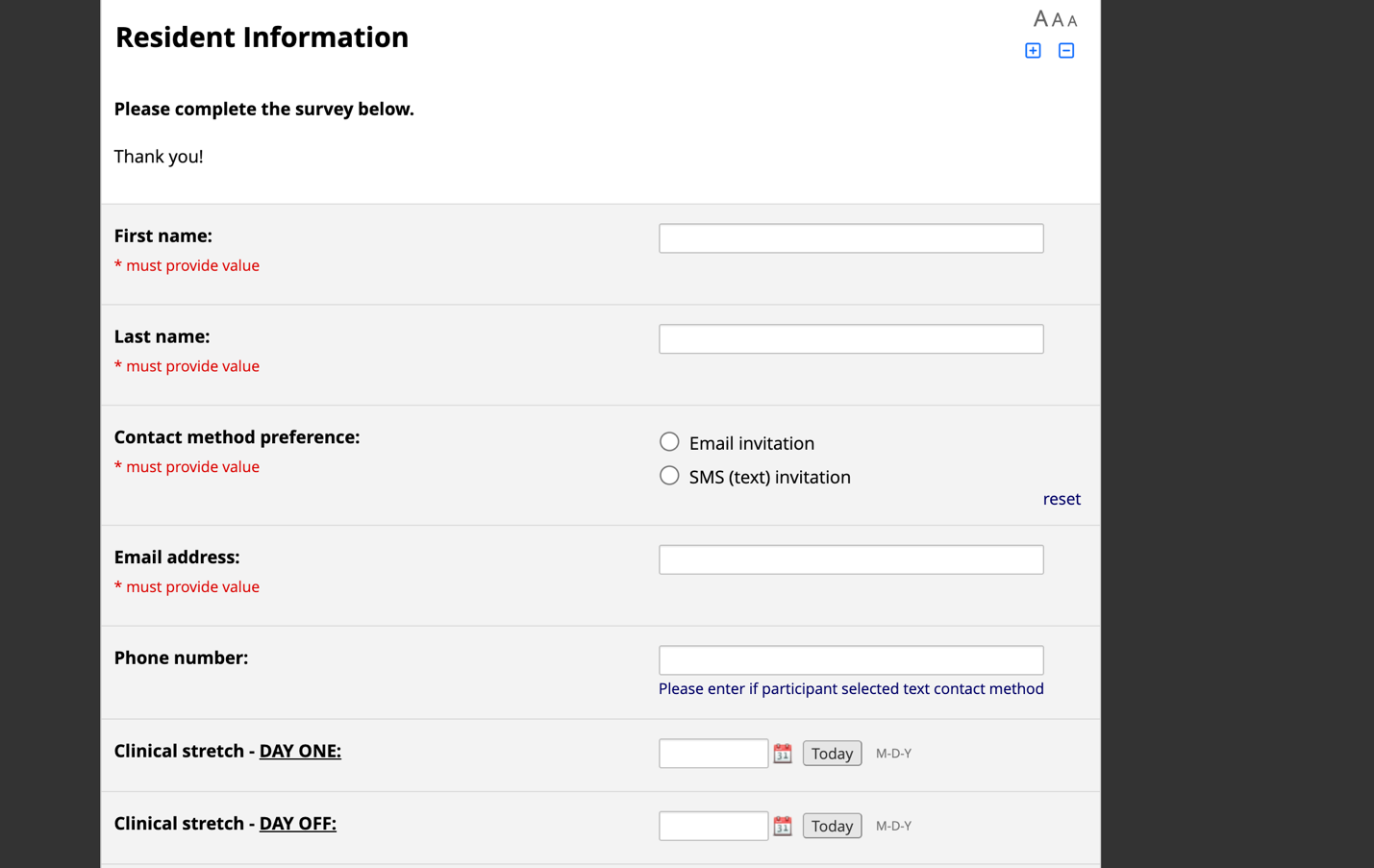

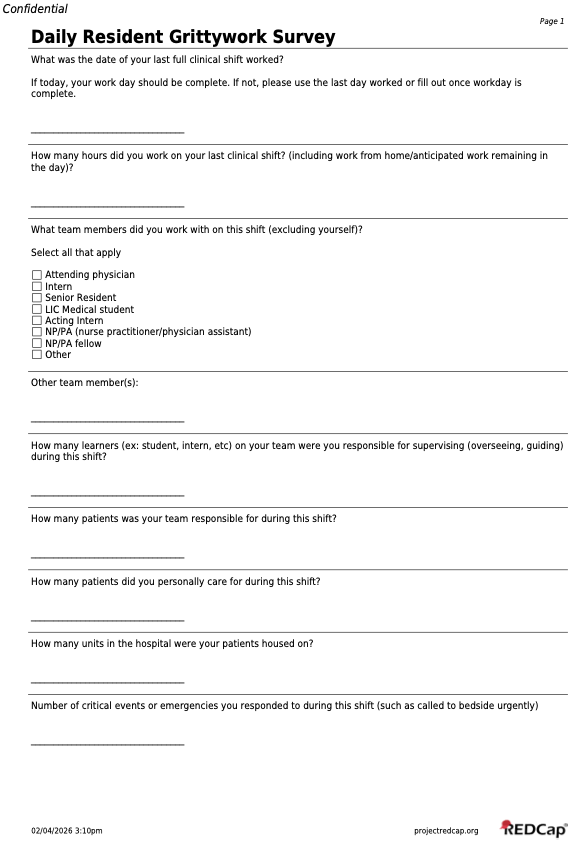

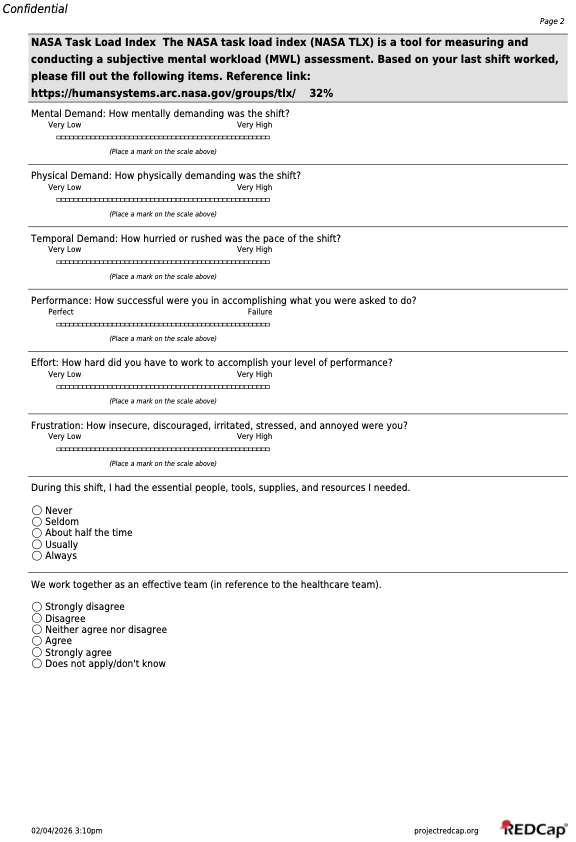

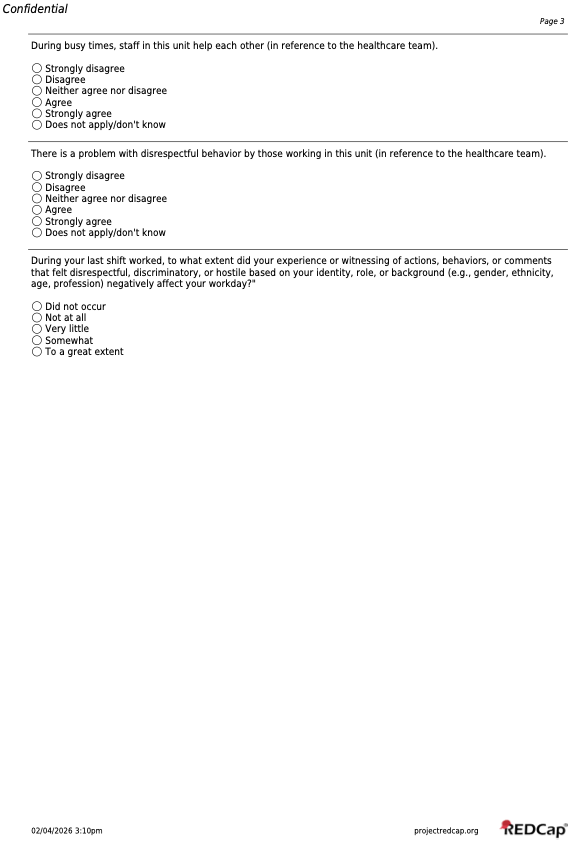

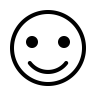

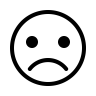

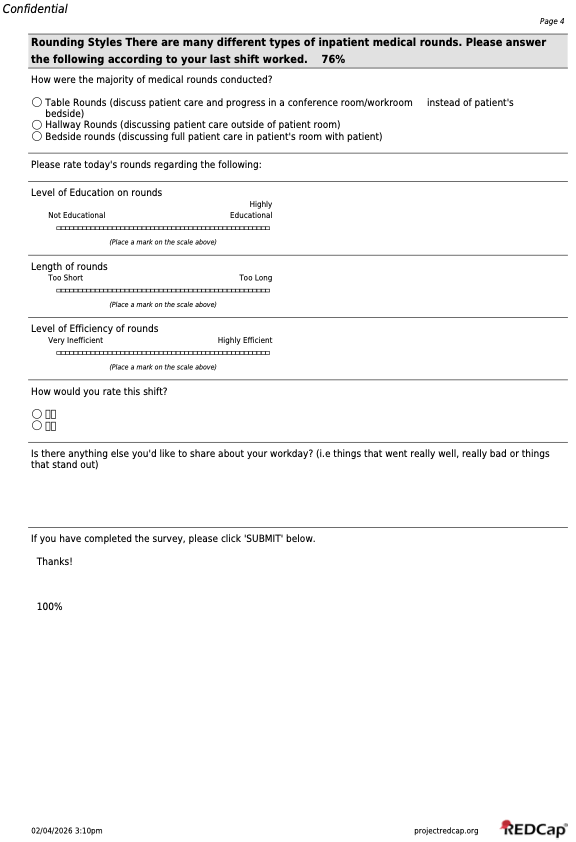

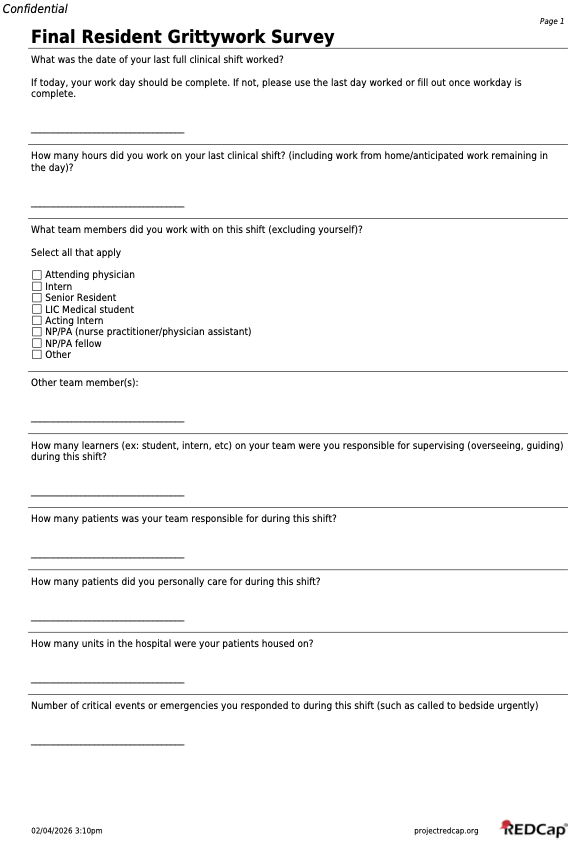

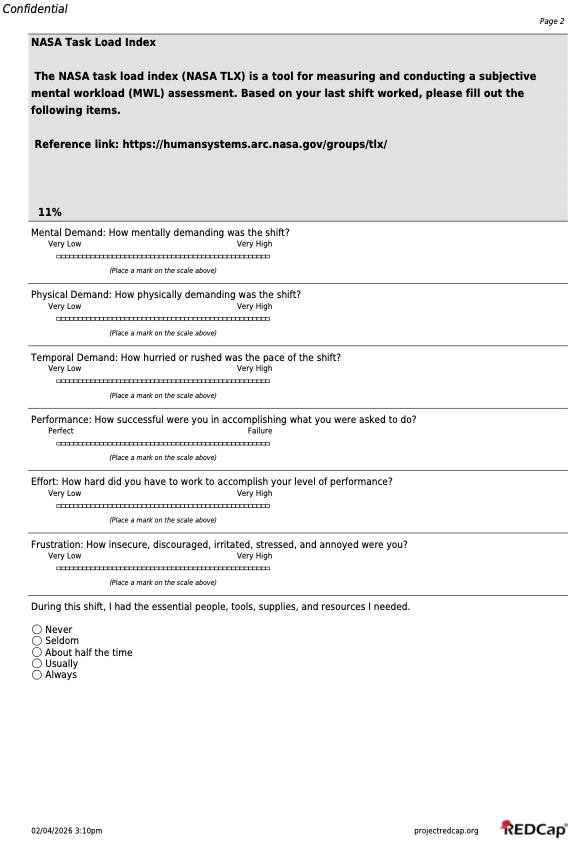

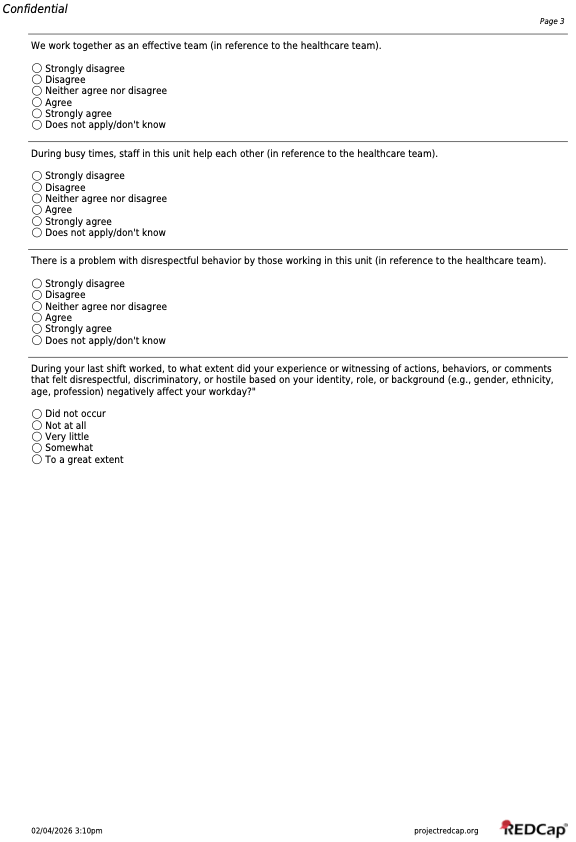

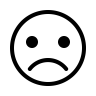

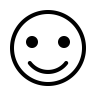

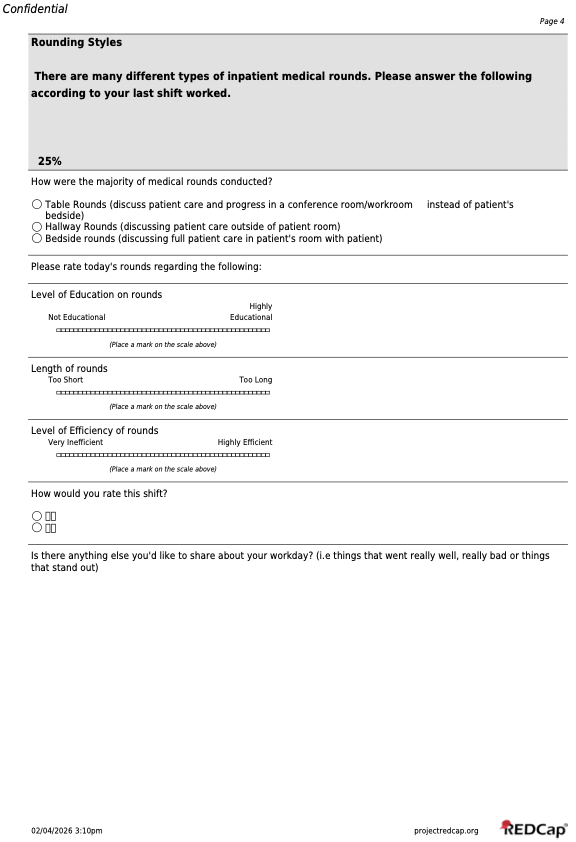

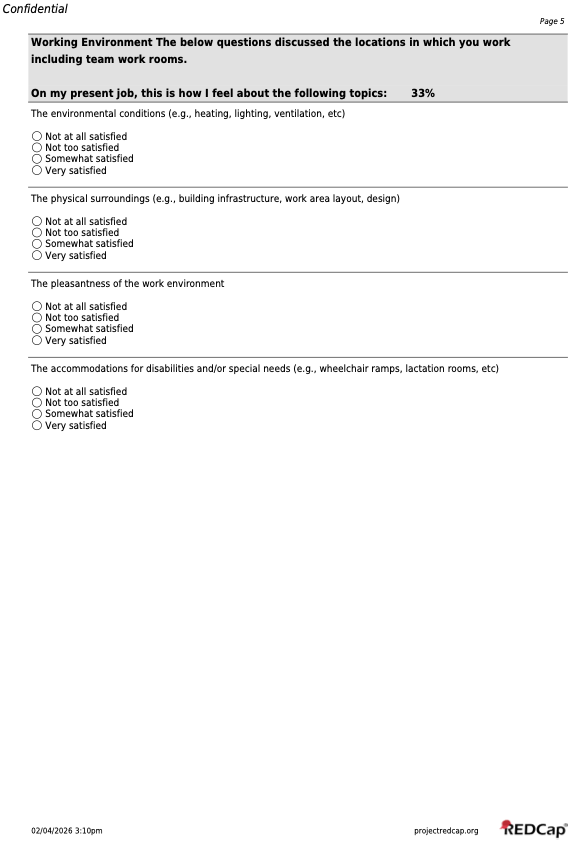

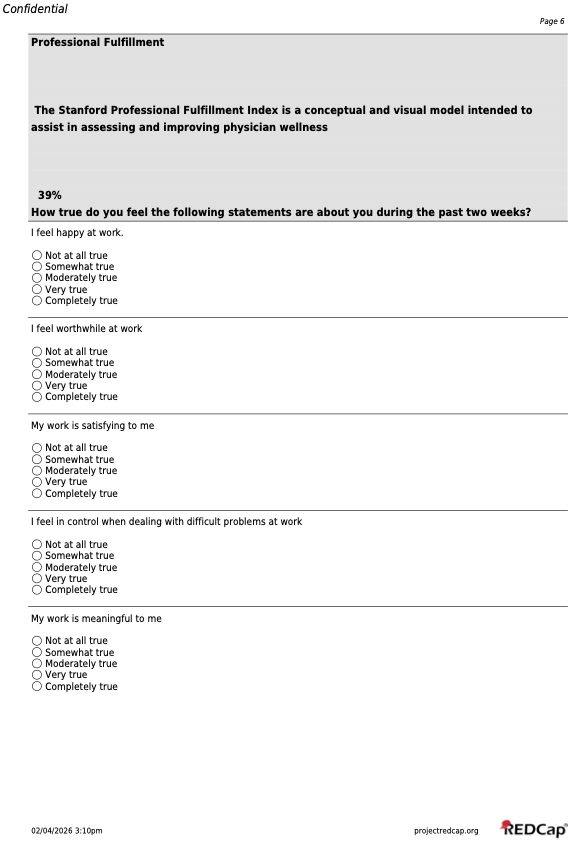

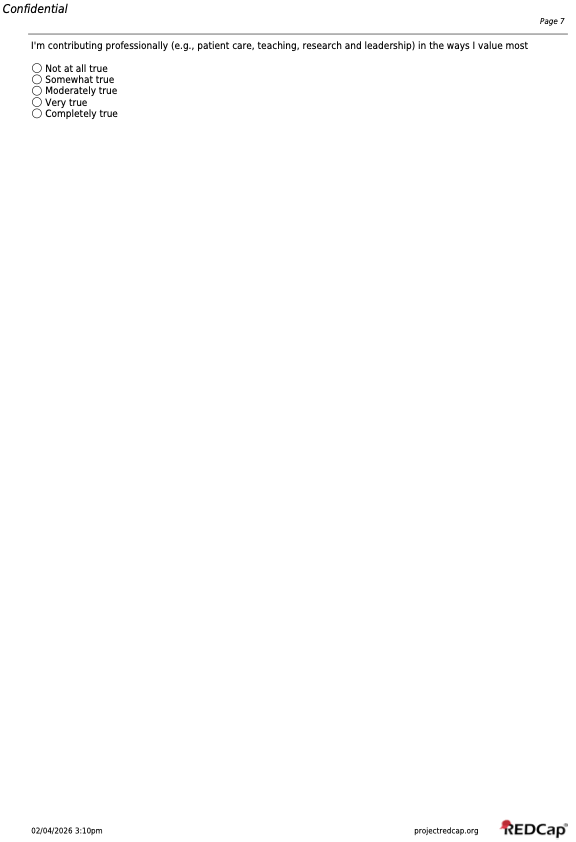

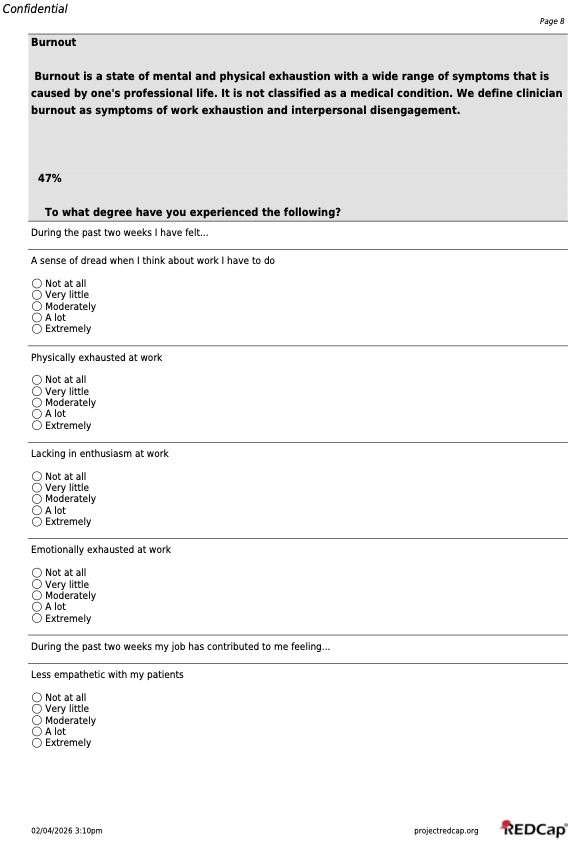

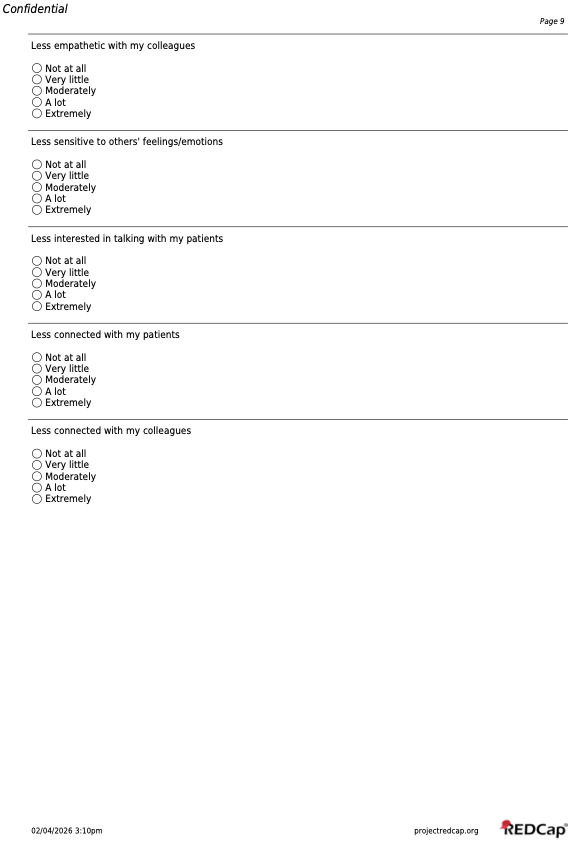

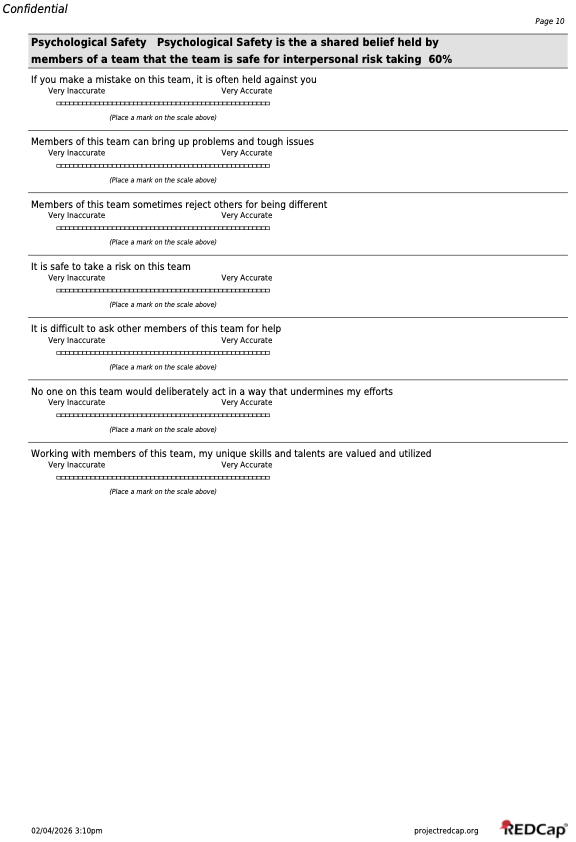

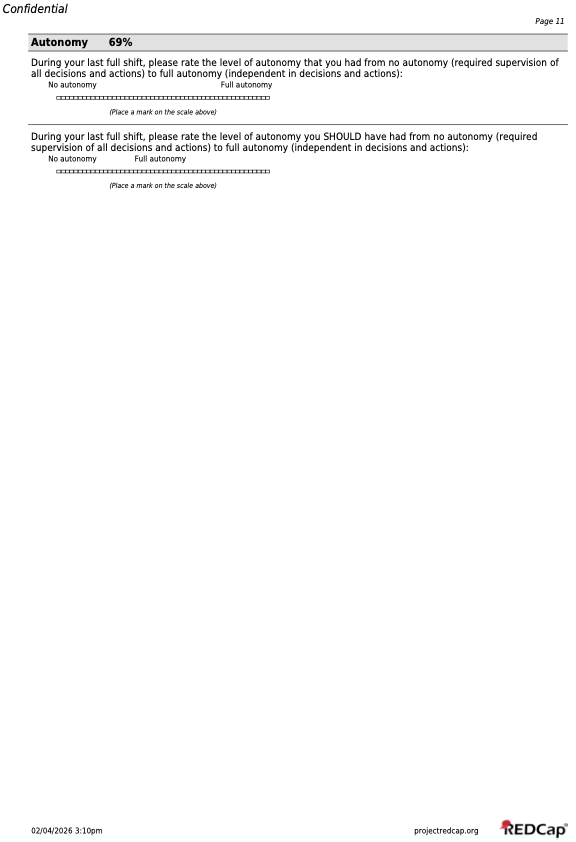

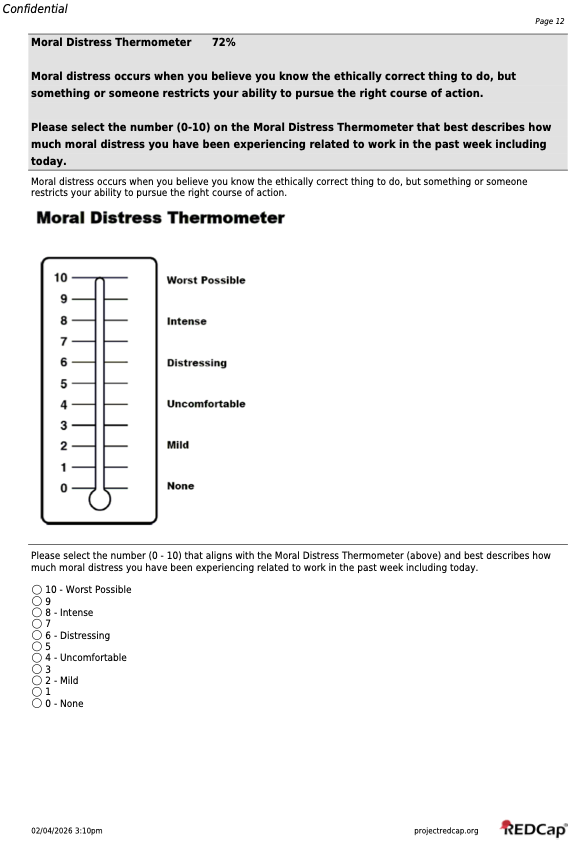

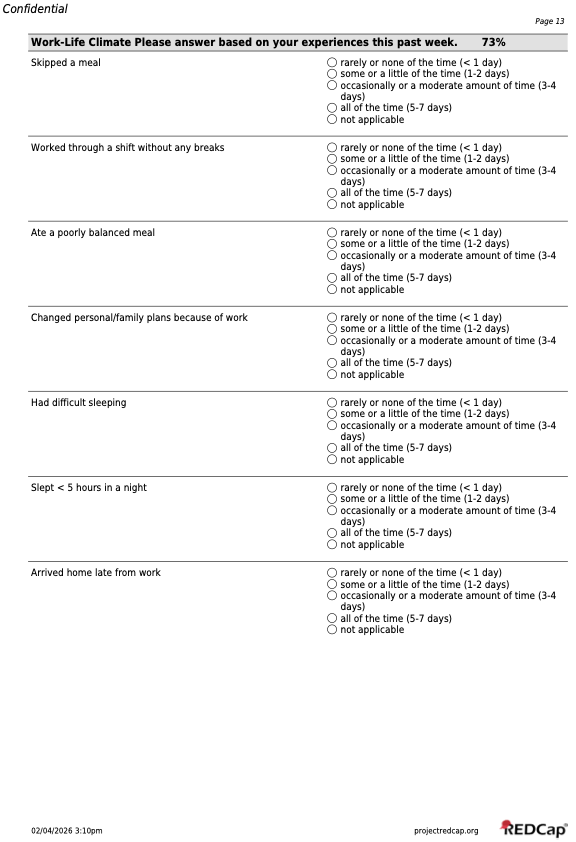

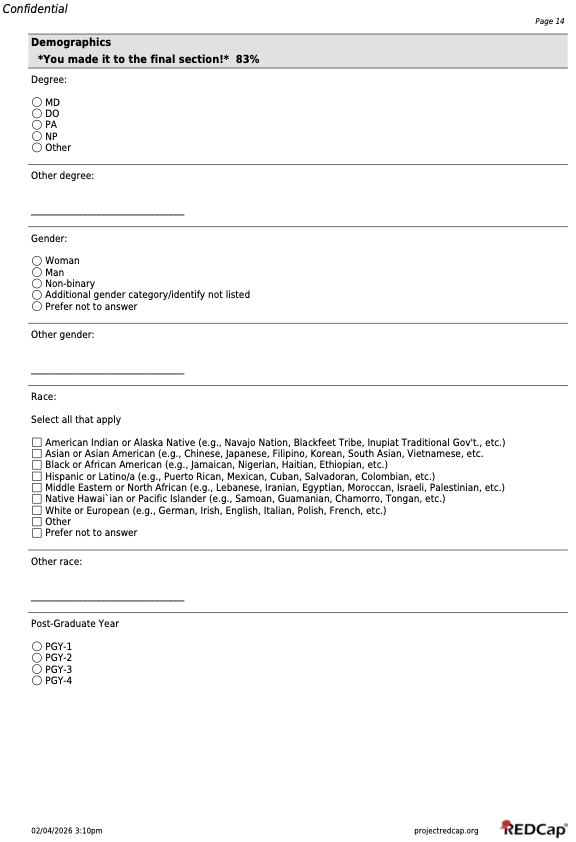

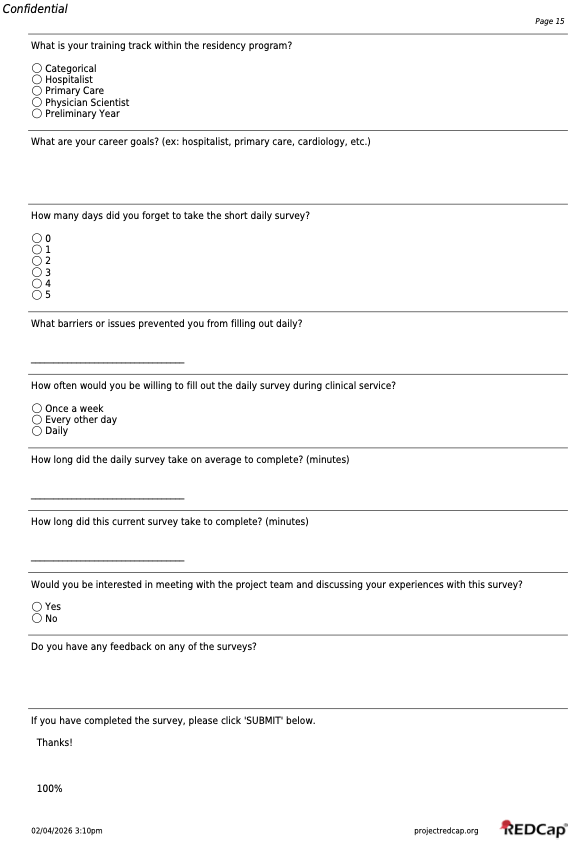

Supplemental Table S1. Post-pilot Survey User Experience Evaluation

| **User Experience Questions** | **Participants (N=28)** |
| --- | --- |
| **How many days did you forget to take the short daily survey?** | **N (%)** |
| 0 | 25 (89) |
| 1 | 2 (7) |
| 2 | 0 (0) |
| 3 | 0 (0) |
| 4 | 0 (0) |
| 5 | 0 (0) |
| Missing | 1 (4) |
| **How long did the daily survey take on average to complete (in minutes)? (Mean ± SD)** | 4.4 ± 1.6 |
| Missing, N | 1 |
| **How long did the current survey take on average to complete (in minutes)? (Mean ± SD)** | 8.6 ± 2.3 |
| Missing, N | 1 |

Supplemental Table S2. Qualitative content analysis of free text responses

Question: Is there anything else you’d like to share about your workday? (i.e. things that went really well, really bad or things that stand out)

| **Domain** | **Working definition** | **Frequency of domain** | **Exemplar comments** |
| --- | --- | --- | --- |
| Team Structure | Number and type of team member | 11 | “Was solo today, lots of work, but good day overall” |
| Care Coordination | Discussions amongst multi-disciplinary team | 9 | “Poor communication from consultants with changing recommendations without notifying team. Led to extra time/secure chats/medication changes clarifying plan at last minute.” |
| Disruptions/Task Switching | Breaks in concentration or flow | 8 | “Two morning admits ruining the flow of rounds once again. Complex patients and coordination of care took too long so I couldn't start on notes till late afternoon but then all the nursing chats prevented me from doing notes and signing out on time.” |
| Procedural Workload | Number and complexity of procedure | 7 | “Also did a paracentesis that leaked so had to address which made shift go long” |
| Admission Timing | Admissions that occur at suboptimal times | 6 | “Admission in morning once again messed up workflow. Rounds lasted too long” |
| Length of Rounds | Time spent on rounding | 6 | “Too many things came up that prolonged rounds” |
| Hours Working | Total number of hours worked (including outside hospital) | 6 | “Two morning admits ruining the flow of rounds once again. Complex patients and coordination of care took too long so I couldn't start on notes till late afternoon but then all the nursing chats prevented me from doing notes and signing out on time.” |
| Educational Experience | Receiving education to assist in growth/development | 6 | “There were a lot of interesting patients so despite being the first day, which can be very hectic, it was enjoyable from the educational perspective.” |
| Number of Discharges | Number of discharges in a day | 5 | “Intern alone day with 4 discharges and 1 admit” |
| Difficult Interactions | Conversations and encounters that are hard to navigate emotionally and professionally | 5 | “Patient very ill/ AMS and their family has zero trust in the medical system, a lot of mental energy thinking of how to make them feel safer and talking to them, but not always feeling successful doing so” |
| Secure Chats | Communication by secure chat | 5 | It is super frustrating that we cannot organize chat by patient. Often, sometimes you get chats and are pulled into multiple different directions for a different patients or you have several chats going about the same patient and it's hard to keep things straight. |
| Team Familiarity | Team members knowledge of the medical team's patients | 4 | “first day with new attending, but able to get everyone on the same page pretty quickly” |
| Overnight Admits | Number of new patients to team overnight | 3 | “Lots of new patients (3 overnight, 2 admits before 8 am), lots of complex care coordination, tricky goals of care discussions and one almost AMA discharge. ” |
| Efficiency of Rounds | Minimizing delays and redundancy during rounds | 3 | “Team tried table rounds as new approach today, significantly shortened rounds and increased efficiency of rounds (more orders placed during rounds, data reviewed much faster)” |
| Number of Admissions | Number of admissions | 3 | “intern alone day. 4 new patients. 3 discharges, and a procedure. Felt like a lot. going home with a few discharge summaries to write, but feeling good about the work done.” |
| Poor Communication | Messages sent/received in a way that led to confusion or error or not received at all | 3 | “Made much worse by one very complex discharge and unresponsive ESD from consulting team” |
| Patient Acuity | How sick is a patient, more urgent evaluation needed | 3 | “Rounding interrupted by 2 emergencies- one right after another. Second was code stroke” |
| Discharge Complexity | Difficulty ensuring safe and effective transition of care | 2 | “A lot of discharges today that were complicated” |
| Resources for Work | Supplies, materials, technology needed to perform job | 2 | “Good shift, four overnight admits and two discharges on procedure service so it was busy. Need more access to computers on rounds” |
| Team Dynamics | Culture and interactions amongst team members, not just who is on team | 2 | “I have an all around above average/fantastic team with great vibes. Feel really lucky this week.” |
| Technology Capability | Is the technology available able to do what you need it to do | 2 | “It is super frustrating that we cannot organize chat by patient. Often, sometimes you get chats and are pulled into multiple different directions for a different patients or you have several chats going about the same patient and it's hard to keep things straight.” |
| Time at Bedside | Time spent with patient or family at bedside | 2 | “Lengthy goals of care conversation (45 min) after signout.” |
| Professional Fulfillment | Feeling work matters and fits who you are | 2 | “Long day, but spent a lot of time at bedside particularly doing a palliative care/GOC discussion, and transitioned a patient to comfort care in a way that felt very in line with her goals, which somehow made the day feel better even though it was longer than normal. I guess it felt meaningful” |
| Work not role aligned | Performing tasks not felt within your job duties | 2 | “Once again rounds way too long and the amount of social work and care coordination chats all day long was incessant. Again wish I could just focus on medicine and have other people take care of the social work type things.” |
| High Patient Volume | Number of patients cared for above typical expectations | 2 | “There is a rule about bounce back admissions that caused us to have to carry 13 patients with the usual cap of 12 and get an admission on our ed day. Triage frustratingly forced us to take an admission because I had one day of overlap with this patient when I joined the team, but they had had a patient directed discharge on my first day getting to know the team, so there was really no continuity. It was poor triaging with no room for conversation.” |
| Patient Complexity | Number of patient comorbidities, health status, active medical problems | 2 | “Two morning admits ruining the flow of rounds once again. Complex patients and coordination of care took too long so I couldn't start on notes till late afternoon but then all the nursing chats prevented me from doing notes and signing out on time.” |
| Unaccounted Work | Work that does not get recorded or acknowledged in system | 1 | “First day of new attending and second day of new team with early AM admission and learner doing their very first admission which significantly disrupts work flow. Had additional patient workup in ED during rounds who ultimately decided to decline admission but significant time and effort dedicated to taking care of this patient. This patient was not counted as 12th patient of the day. Finally, lengthy bedside rounding on a single one of our patients taking ~45 min during rounds.” |
| Social Complexity | Factors that influence care outside the hospital | 1 | “7 new patients today, significant social complexity with multiple patients and frustration from both patients and medical team about lack of resources to help secure outpatient follow up” |
| Limited Resources | Not having enough resources to take care of patients | 1 | “7 new patients today, significant social complexity with multiple patients and frustration from both patients and medical team about lack of resources to help secure outpatient follow up” |
| Poor Educational Experience | Unsatisfactory learning environment or event | 1 | “Hard shift and was told I would get a procedure but pt refused having a learner do it” |
| Teaching | Teaching that you provide to team members | 1 | “First day of new attending and second day of new team with early AM admission and learner doing their very first admission which significantly disrupts work flow. Had additional patient workup in ED during rounds who ultimately decided to decline admission but significant time and effort dedicated to taking care of this patient. This patient was not counted as 12th patient of the day. Finally, lengthy bedside rounding on a single one of our patients taking ~45 min during rounds.” |
| Micro/Macroaggression | A statement, action, or incident regarded as an instance of indirect, subtle, or unintentional discrimination against members of a marginalized group such as a racial or ethnic minority. | 1 | “2nd workday intern alone with full team. Had patient refer to medical student in disrespectful manner, I didn't do enough to respond and correct in the moment. learning experience for me.” |
| No Breaks | Lack of breaks in workflow | 1 | “Nonstop grind without breaks. Lots of care coordination and social work again. Also a scheduling mishap occurred and there was no APP fellow today. So I worked two days in a row as a solo resident which is never ideal.” |

Supplemental material 3. Survey Free Text Responses

**Daily Surveys:**

**Other team member(s):**

- 2 MD-PhD students
- Interprofessional team members x3
- Other staff
- Pharmacy students x2
- Visiting foreign physician x4

**Weekly Survey:**

**What are your career goals? (ex: hospitalist, primary care, cardiology, etc.)**

- academic medicine and research
- Cardiology x3
- Community GI
- Geriatrics
- GI
- Heme onc x3
- Hospitalist x7
- Hospitalist vs cardiology
- Nephrology
- Physician scientist
- Primary care x2
- Renal/GI
- Rural hospitalist
- split hospitalist and palliative

**What barriers or issues prevented you from filling out daily? (N = 2)**

- I forgot the date it started
- Work - busy and forgot

**Do you have any feedback on any of the surveys?**

- Appropriate length, would not make any longer or would likely have not completed on several days
- some questions asked to distinguish between "did not occur" and "not at all" and im not really sure what the intended difference between those answers is.
- Thanks for doing this research!
- The direct patient care I did question was a little confusing and I think I filled it out wrong
- The surveys were very easy to complete!!

Supplemental Table S3. NASA-TLX scores by shift rating

| **Shift rating** | **NASA-TLX (mean** ± **SD)** |
| --- | --- |
| Happy emoji (N = 117) | 46 ± 11 |
| Sad emoji (N = 48) | 62 ± 9 |

P <0.0001

Supplemental Figure S1. Box plots for NASA-TLX scores by shift rating

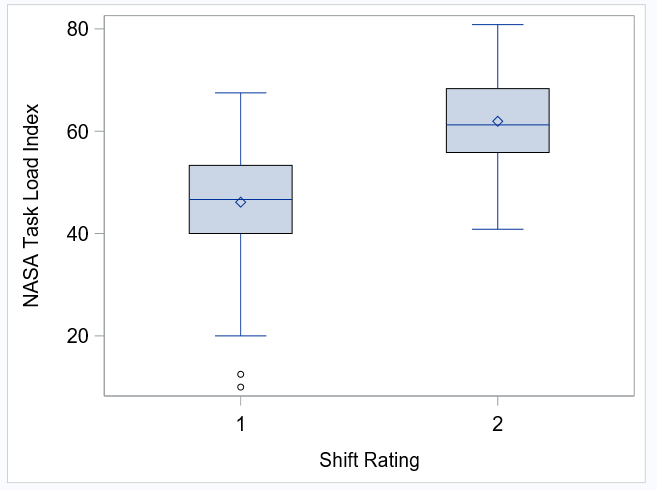

Shift rating 1 corresponds to “happy face” emoji and shift rating 2 corresponds to “sad face emoji”.

Supplemental Table S4. Rounding length score by shift rating

| **Shift rating** | **Length of rounds (mean** ± **SD)** |
| --- | --- |
| Happy emoji (N = 117) | 6 ± 1 |
| Sad emoji (N = 48) | 7 ± 2 |

Correlation coefficient estimate 0.35 (95% CI 0.21, 0.48), P<0.0001.

Supplemental Figure S2. Box plots for rounding length score by shift rating

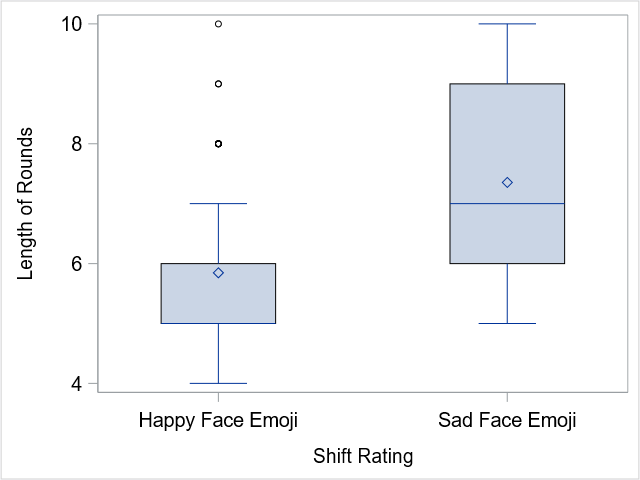

Supplemental Figure S3. Scatter plot for correlation between length of rounds score and NASA-TLX score

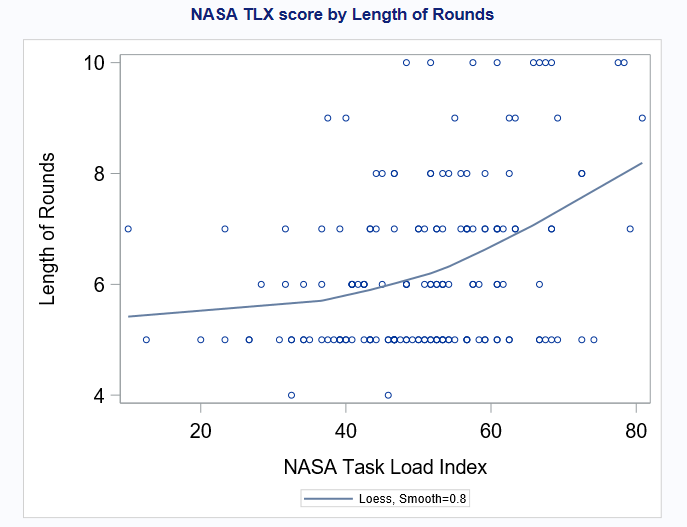
